# A multidimensional Mendelian randomization study on the impact of gut dysbiosis on chronic diseases and human longevity

**DOI:** 10.1101/2021.08.20.21262026

**Authors:** Éloi Gagnon, Patricia L. Mitchell, Hasanga Manikpurage, Erik Abner, Nele Taba, Tõnu Esko, Nooshin Ghodsian, Sébastien Thériault, Patrick Mathieu, Benoit J. Arsenault

## Abstract

Alterations of the gut microbiota, often referred to as gut dysbiosis, have been associated with several chronic diseases and longevity in pre-clinical models as well as in observational studies. Whether these relationships underlie causal associations in humans remains to be established. We aimed to determine whether gut dysbiosis influences the risk of chronic diseases and longevity using a comprehensive 2-Sample Mendelian randomization (2SMR) approach. We included as exposures inflammatory bowel disease (IBD) as a human model of gut dysbiosis, 11 gut-associated metabolites and pathways and 48 microbial taxa. Study outcomes included eight chronic diseases previously linked with gut dysbiosis using observational studies (Alzheimer’s disease, depression, type 2 diabetes, non-alcoholic fatty liver disease, coronary artery disease (CAD), stroke, osteoporosis and chronic kidney disease) as well as parental longevity and life expectancy. Neither IBD, nor gut-associated metabolites were causally associated with chronic disease or lifespan. After multiple testing correction for 582 tests, no microbial taxa-chronic disease associations remained significant. After robustness analyses and multivariate MR to correct for body mass index and alcohol intake on all 42 nominally significant causal relationships, four associations remained. Altogether, results of this multidimensional Mendelian randomization study suggest that gut dysbiosis has little impact on chronic diseases and human longevity and that previous documented associations may not underly causal relationships. Studies with larger sample sizes and more optimal taxonomic discrimination may ultimately be required to determine whether the human gut microbiota plays a causal role in the etiology of chronic diseases and longevity.

## Introduction

The human gut microbiota is the microbial symbiotic organ residing in the gut. It is involved in key metabolic and immunological processes including host immunity, food digestion, intestinal endocrine function and intestinal permeability (Lynch and Pedersen 2016). Several observational studies revealed that alterations of the gut microbiota, often referred to as gut dysbiosis, are associated with a wide range of diseases and metabolic conditions (Hooks and O’Malley 2017). In individuals with gut dysbiosis, dysfunction of the gut mucosal barrier (“leaky gut”) may lead to an increased bacterial translocation through the mucosal cell line and into the blood, thereby affecting the homeostasis of other organs and promoting disease development and progression (Anhê et al. 2020; Konturek et al. 2018).

Inflammatory bowel disease (IBD), comprising ulcerative colitis and Crohn’s disease, is the most frequently reported disease unequivocally associated with gut microbiota changes and gut inflammation (Schippa and Conte 2014). It involves chronic inflammation of the digestive tract due to aberrant immune response, possibly mediated by dysbiosis. Many studies have reported that the composition of microbiota in IBD is altered compared to healthy subjects (Andoh et al. 2007; Fujimoto et al. 2013; Nishino et al. 2018; Takahashi et al. 2016). Gut dysbiosis could contribute to inflammation and immune responses leading to IBD and this inflammatory environment could in turn promote dysbiosis (Sartor and Wu 2017). Results of several prospective observational studies suggested that patients with IBD may be at increased risk of multiple chronic diseases such as cardiovascular disease (Cainzos-Achirica et al. 2020), respiratory disease, arthritis, liver conditions, kidney failure (Xu 2018), osteoporosis and depression (Bernstein et al. 2019), consistent with the view that dysbiosis exerts systemic negative effects promoting human diseases.

The systemic effects of gut dysbiosis are partly mediated through by-products of the microbiome. These microbial metabolites can reach the peripheral circulation via the portal vein (den Besten et al. 2013), or diffuse readily and be taken up by the gut mucosa (Flint 2016), where they can reach organs and act as substrates or signaling molecules. This bidirectional crosstalk between the gut microbiota and different organs occurs via the gut-liver axis (Albillos, Gottardi, and Rescigno 2020), the gut-brain axis (Carabotti et al. 2015), the gut-bone axis (Villa, Ward, and Comelli 2017), the gut-kidney axis (Evenepoel, Poesen, and Meijers 2017), the gut-lung axis (Marsland, Trompette, and Gollwitzer 2015) and the gut-heart axis (Bartolomaeus, McParland, and Wilck 2020). Specific classes of microbiota-derived metabolites, notably short-chain fatty acids (De Vadder et al. 2014), branched-chain amino acids (Arany and Neinast 2018), trimethylamine N-oxide (Shan et al. 2017) and indole (Beaumont et al. 2018) have been strongly implicated in the pathogenesis of metabolic disorders (Agus, Clément, and Sokol 2020), lifespan (Wilmanski et al. 2021), neurological and cardiovascular diseases (Martinez, Leone, and Chang 2017).

Over the past few years, the gut microbiota emerged as a therapeutic target of great interest to prevent and/or treat chronic diseases and improve human lifespan and healthspan. An overwhelming amount of supportive evidence from pre-clinical models contributed to the widely-accepted view that a large number of diseases and pathological processes could be influenced by the microbiome, from early metabolic perturbations to full-blown diseases and premature mortality. Fecal transplantation studies in rodents have provided promising results for the treatment of obesity (Pérez-Matute et al. 2020), type 2 diabetes (T2D) (H. Wang et al. 2020), depression and chronic stress (Langgartner et al. 2018), liver injury (Y. Liu et al. 2021), myocarditis (Hu et al. 2019) and aging (Chen et al. 2020). Human microbiota-associated (HMA) studies, consisting of the transplantation of feces from human patients into germ-free mice while control mice receive feces from healthy humans, further supported these associations.

A systematic review conducted in 2019 on the HMA method to study the impact of the microbiota on chronic diseases reported that 95% of such studies (36/38) concluded that fecal transplantation from a sick human donor resulted in at least one worsened symptom compared to healthy controls (Walter et al. 2020). This finding was deemed “implausible” by the authors of this systematic review (Walter et al. 2020). According to Walter et al., in the vast majority of cases, these studies lacked adequate replication and they had statistical and methodological flaws that artificially inflated the odds of obtaining positive findings (Walter et al. 2020). Together with the “file-drawer effect” (whereby positive studies are more likely to be published compared to negative studies), these caveats may distort the odds of translating findings from pre-clinical models into microbiota-targeting therapies to prevent or treat human diseases. Observational studies in humans with various diseases have identified relevant differences in intestinal microbiota composition (Shreiner, Kao, and Young 2015). However, they are subject to biases such as reverse causality and confounding (through unmeasured confounders) and cannot, by design, assess causality. Obesity, pharmacotherapy, diet, alcohol intake and many other factors appear to be important confounders in the microbiota-health relationships (Vujkovic-Cvijin et al. 2020). Given these limitations, Walter et al. suggested that novel and innovative methods such as Mendelian randomization (MR) should be used to investigate the causal role of the gut microbiota in human disease etiology.

MR is an epidemiological approach that is not subject to many of the biases of observational studies such as reverse causality or confounding. It has the potential to evaluate multiple microbiota-health relationships all at once in a hypothesis-free manner. Briefly, MR uses genetic variants strongly associated with an exposure (features of human gut dysbiosis) to infer causality with an outcome (chronic diseases and human longevity). Twin studies have shown that heritability of the abundance of different bacterial taxa is on average 20%, although some variation exists between taxa (Goodrich et al. 2014; 2016). This is consistent with the view that genes play a non-negligible role in determining gut microbiota composition, making MR a valuable tool to assess the potential causal role of gut dysbiosis in human diseases.

Here, we used a 2-sample MR (2SMR) study design to investigate the potential causal link between three levels of gut dysbiosis-related exposures and eight chronic diseases-related outcomes (coronary artery disease [CAD], T2D, ischemic stroke [IS], nonalcoholic fatty liver disease [NAFLD], chronic kidney disease [CKD], osteoporosis, Alzheimer disease [AD] and depression) and human lifespan (as defined by parental lifespan and living beyond the 90^th^ percentile). In our “multidimensional” MR design, we first used IBD as a human model of gut dysbiosis (Schippa and Conte 2014) to determine its association with chronic diseases and human lifespan using multiple MR methods. Second, we investigated the potential causal association of fecal and blood metabolites associated with gut dysbiosis and disease. Third, we leveraged summary statistics from two large genome-wide association studies (GWAS) of gut microbial signature to investigate the association between genetically predicted gut microbes and taxa abundance with chronic diseases and human longevity.

## Results

The conceptual framework of this MR analysis as well as the datasets used to derive the study exposures and outcomes are presented in Figure 1 and Table 1. Briefly, the objective of this multidimensional MR analysis was to test the hypothesis that the gut microbiome causally impacts chronic diseases and longevity and to provide estimates for each exposure/outcome association. We performed 2SMR on exposures associated with gut dysbiosis and relevant outcomes of human diseases and longevity. We used publicly available genome-wide association study (GWAS) summary statistics to extract 10 disease-related outcomes and human longevity (see Methods) and 60 traits related to the microbiome including IBD as a model of human gut inflammation and dysbiosis, fecal and plasma metabolites associated with the gut microbiota as well as microbial abundance of taxa partly under genetic control. Analyses were restricted to participants from European ancestry except for the study by Kurilshikov et al. 2021. The sample from study exposure and outcome did not overlap. We selected only exposures that had at least three independent (r2 <0.01) genetic instruments at minimum p-value <1e-5 (the threshold differed between exposures depending of the availability of genetic instruments) with mean F statistics >15, resulting in 60 microbiota-related exposures available for MR.

**Figure 1.**
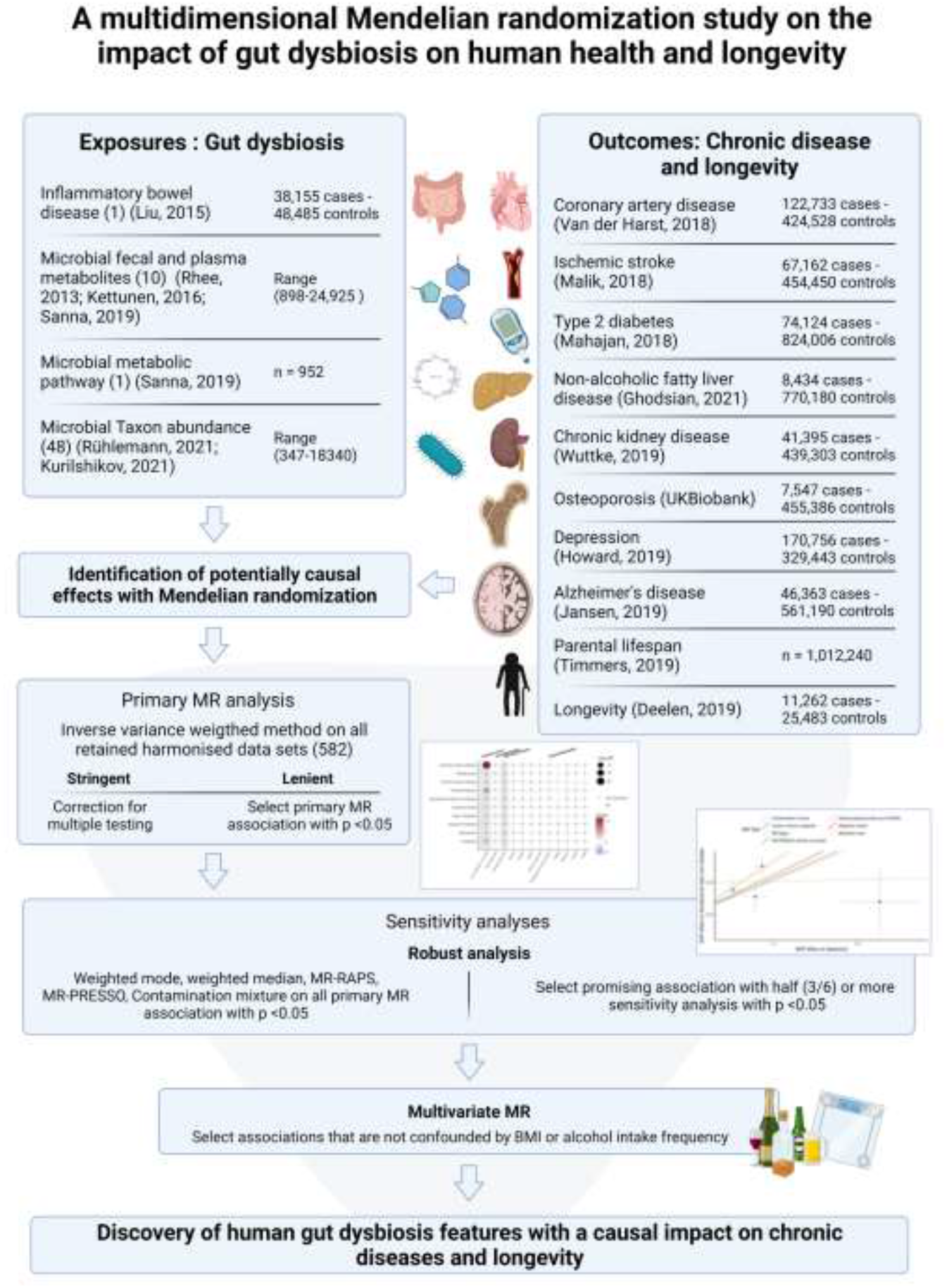
Overview of the multidimensional Mendelian randomization framework used to investigate the association between gut dysbiosis (inflammatory bowel disease, microbial metabolites and microbial taxa abundance) and chronic diseases and longevity.

**Table 1.** Description of cohorts used in this study.

|  | Trait | Cohort of study | Sample size | Populations | (Author, year) | pubmed id |
| --- | --- | --- | --- | --- | --- | --- |
| **Exposures** |  |  |  |  |  |  |
| Diseases | Inflammatory Bowel Disease | only european individuals from BEL1, BEL2, CEDARS, CHOP, GERMAN, NIDDK, WTCCC, NORWEGIAN and SWEDISH. | 38,155 cases - 48,485 controls | European | (Liu et al., 2015) | 26192919 |
| functional microbial pathways | PWY-5022 | LifeLines-DEEP | 952 | European | (Sanna et al., 2019) | 30778224 |
| fecal metabolites | Propionate | LifeLines-DEEP | 898 | European | (Sanna et al., 2019) | 30778224 |
| fecal metabolites | TMAO, indole-3-propionate (IPA), carnitine, betaine and choline | Framingham Heart Study offspring cohort | 2076 | European | (Rhee et al., 2013) | 23823483 |
|  | leucine, isoleucine and valine and acetate | EGCUT, ERF, FTC, FR97, COROGENE, GenMets, HBCS, KORA, LLS, NTR, NFBC, PredictCVD, PROTE, YFS | 24,925 | European | (Kettunen et al., 2016) | 27005778 |
| Microbial Abundance | Microbial relative abundance | PopGen, FoCus, KORA FF4, SHIP | 8,956 | European | (Rühlemann et al., 2021) | 33462482 |
|  | Microbial relative abundance | BSPSPC, CARDIAb, CARDIAw, COPSAC, DanFunD, FGFP, FOCUS, GEM, GenR, KSCS, LLD, METSIM, MIBS, NGRC, NTR, PNP, POPCOL, RS3, SHIP, HCHS/SOL, TwinsUK | 18,34 | Mixed (>72%European) | (Kurilshikov et al., 2021) | 33462485 |
| **Outcomes** |  |  |  |  |  |  |
| Chronic diseases | Coronary artery disease | CARDIoGRAMplusC4D and UK Biobank | 122,733 cases - 424,528 controls | European | (Van der Harst, 2018) | 29212778 |
|  | Ischemic stroke | MEGASTROKE consortium | 67,162 cases - 454,450 controls | European | (Malik, 2018) | 29531354 |
|  | Type 2 diabetes | DIAGRAM consortium | 74,124 cases -<br>824,006 controls | European | (Mahajan,<br>2018) | 30297969 |
|  | Non-alcoholic fatty<br>liver disease | eMERGE, UK Biobank, Estonian Biobank and<br>FinnGen | 8434 cases -<br>770,180 controls | European | (Ghodsian,<br>2021) | unpublished |
|  | Chronic kidney disease | CKDGen consortium | 41,395 cases -<br>439,303 controls | European | (Wuttke,<br>2019) | 31152163 |
|  | Osteoporosis | UK Biobank | 7547 cases -<br>455,386 controls | European | UKB | PheCode_743.1<br>in the UKB |
|  | Depression | PGC and UK Biobank | 170,756 cases -<br>329,443 controls | European | (Howard,<br>2019) | 30718901 |
|  | Alzheimer's disease | PGC-ALZ, IGAP, ADSP and UK Biobank | 246,363 cases -<br>561,190 controls | European | (Jansen,<br>2019) | 30617256 |
| Lifespan | Parental lifespan | LifeGen and UK Biobank | n = 1012,240 | European | (Timmers,<br>2019) | 30642433 |
|  | Longevity | 100-plus/LASA/ADC, AGES, CEPH, CHS, DKLS,<br>FHS, GEHA, HRS, LLFS LLS, Longevity, MrOS,<br>Newcastle 85 +, RS, SOF, Vitality 90 +, | 11,262 cases -<br>25,483 controls | European | (Deelen,<br>2019) | 31413261 |

### Effect of inflammatory bowel diseases on chronic diseases and longevity

Gut dysbiosis is a hallmark of IBD (Kostic, Xavier, and Gevers 2014) and the presence of IBD influences the microbial signature (Ni et al. 2017). Using an IBD GWAS of 38,155 cases and 48,485 controls (J. Z. Liu et al. 2015a), we first calculated genetic correlation (R_G_) with IBD and the 10 outcomes under investigation (Figure 2a). IBD had a weak to modest genetic correlation with all of the outcomes with the majority not significantly different from zero. IBD had a weak positive R_G_ with depression and a weak negative R_G_ with parental lifespan and CKD. We then selected all independent (r2≤0.001) genome-wide significant single nucleotide polymorphisms (SNPs) (p-value<5e-8) as genetic instruments and performed an Inverse Variance Weighted (IVW)-MR linking IBD with all 10 outcomes. IBD had weak effect on all 10 outcomes under investigation. The only association that reached nominal statistical significance was the one linking IBD to NAFLD (Figure 2b). Each log odds ratio (OR) of IBD increased the risk of NAFLD by 4% (OR=1.04 (1.00-1.08), p=0.02). Robust MR methods were consistent with the absence of causal association for all associations (Supplementary Table 1). Single SNP MR analyses were consistent with a high degree of heterogeneity among the Wald ratios of individual genetic instruments with the majority of IBD SNPs not being associated with diseases (Supplementary Figure 1A-J). Altogether these analyses do not support a causal role of IBD in the etiology of chronic diseases and longevity.

**Figure 2.**
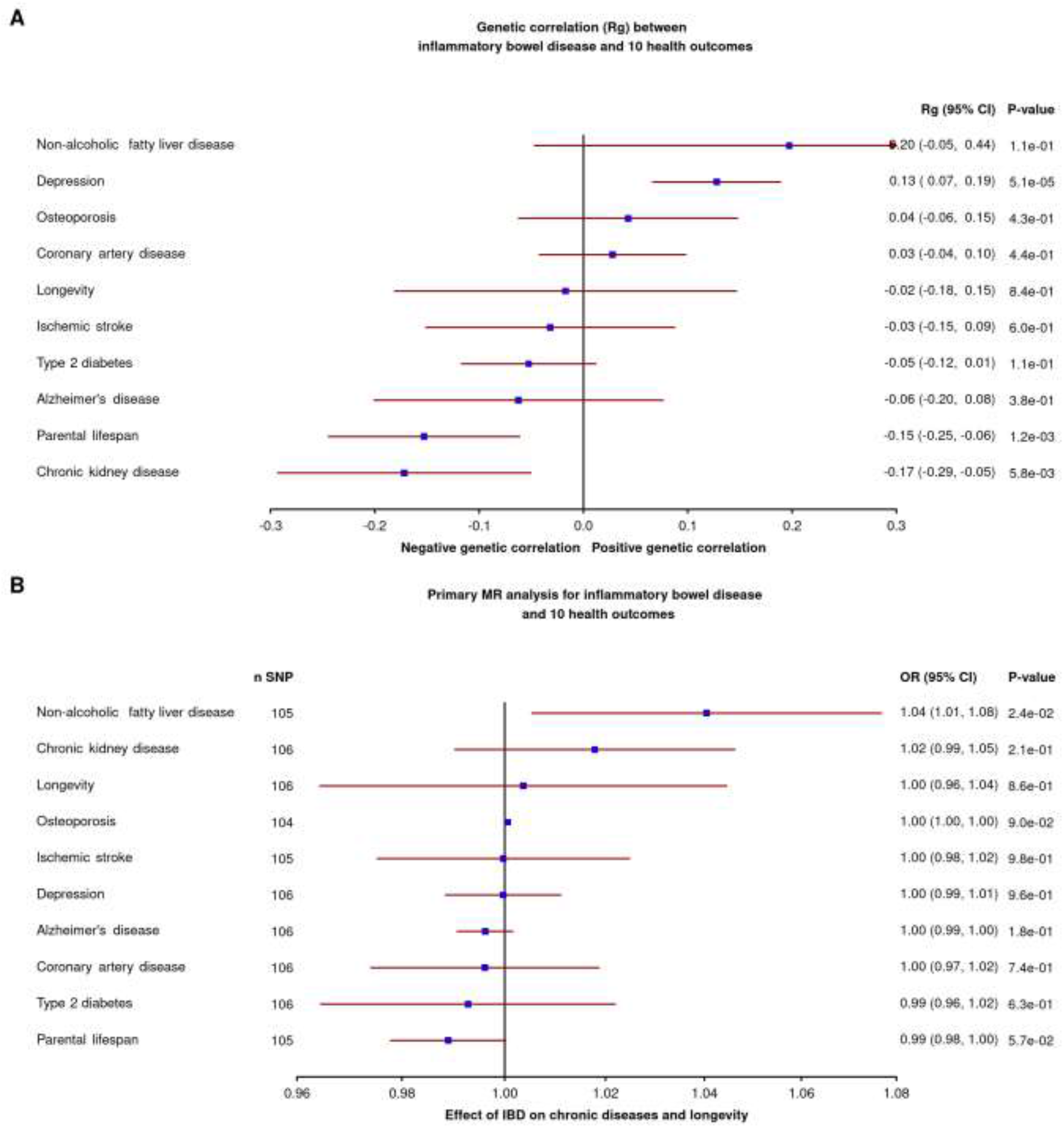
Forest plot of the association of inflammatory bowel disease with all 10 health outcomes. A) Genetic correlation B) MR causal estimates with inverse variance weighted multiplicative random effect.

### Effect of gut microbiome-related metabolites on chronic diseases and longevity

We next sought to determine whether blood metabolites associated with the gut microbiota and fecal microbial metabolites or their functional pathways could influence chronic diseases and longevity. We selected genetic instruments for fecal propionate, a gut metabolite linked with T2D in a recent MR study, from a GWAS of 898 participants (Sanna et al., 2019). We also included 9 plasma metabolites: indole-3-propionate, trimethylamine N-oxide and some of its precursors, carnitine, betaine and choline (Rhee et al. 2013), branched chain amino acids (leucine, isoleucine and valine) as well as the short chain fatty acid acetate (Kettunen et al. 2016). We finally included the microbial pathway involved in 4-aminobutanoate (GABA) degradation (PWY-5022 pathway) acting as a proxy for butyrate production by the gut (Sanna et al., 2019). We selected independent (r2≤0.01) SNPs (for all studies: p-value<1e-5; except Kettunen et al.: p-value<1e-6) as genetic instruments. We performed IVW-MR for each of the health outcomes under study (Figure 3). A total of 104 exposure-outcomes associations were tested. Nine passed a nominal p-value significance threshold of 0.05 (including the propionate-T2D association) but none of the gut microbiota metabolites were associated with chronic diseases and longevity after multiple testing correction. Figure 3 also reports the association of LDL cholesterol, used as positive control with the outcomes of interest. As expected, LDL cholesterol was strongly associated with cardiovascular diseases (OR = 0.43 95% CI 0.36-0.50, P=5.22e-32) and longevity (Beta = -0.11 95% CI -0.15 -0.075, P= 3.94e-09)

**Figure 3.**
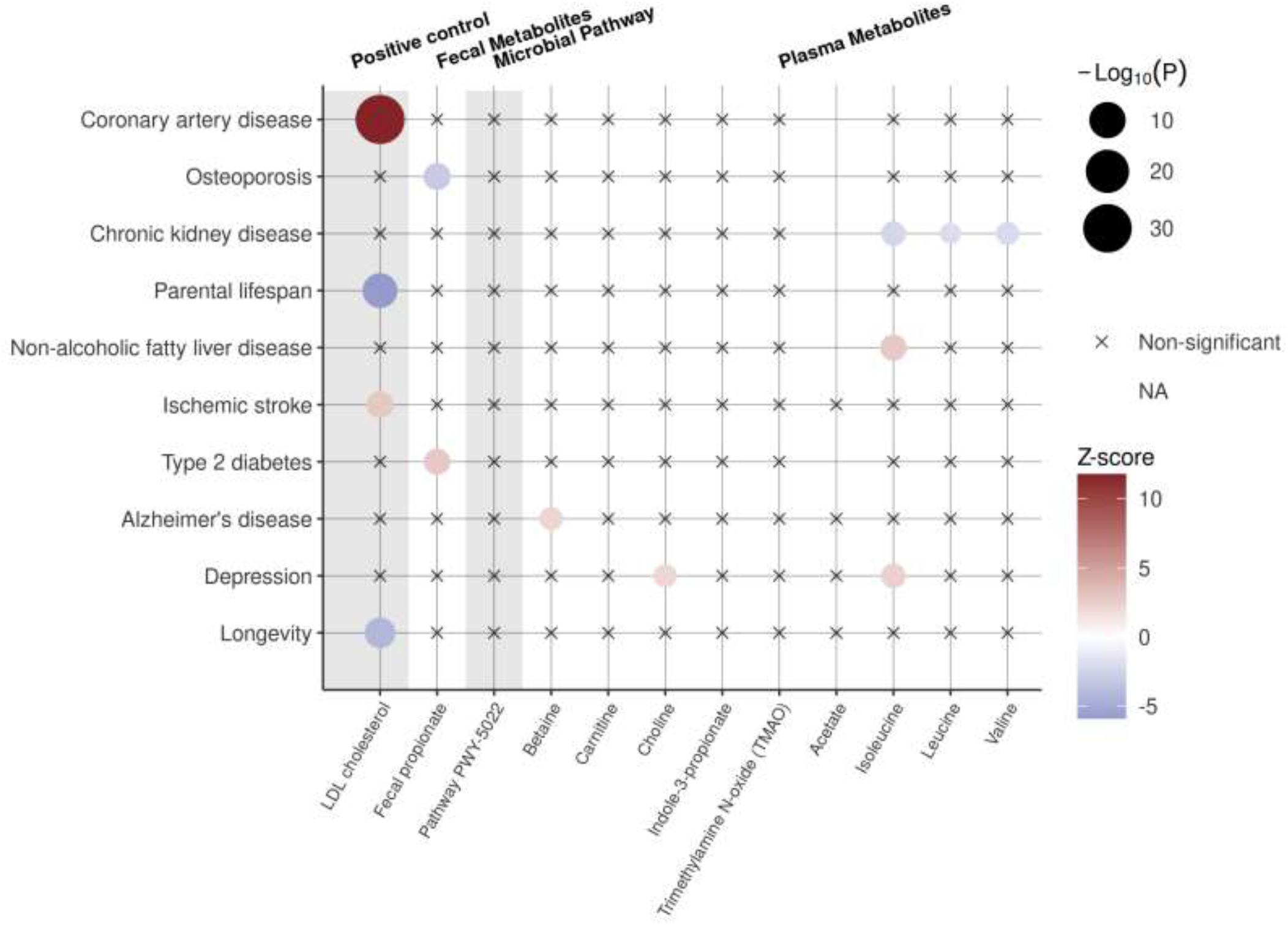
Balloon plot of the association of microbial fecal metabolites, microbial pathway and plasma metabolites with all 10 health outcomes. Associations with LDL cholesterol are added as positive controls. Non-available (NA) associations stem from a lack of overlapping SNPs or proxies between exposure and outcome resulting in fewer than three genetic instruments in the harmonized data set. Non-significant associations (p-value > 0.05) are depicted with crosses.

### Effect of gut microbial abundance on chronic diseases and longevity

We finally explored the impact of different taxa abundance on health-related outcomes. We first identified 4 recent GWAS on gut microbe abundance with available summary statistics (Kurilshikov et al. 2021; Lopera-Maya et al. 2020; Qin et al. 2020; Rühlemann et al. 2021). We then filtered all microbiota quantitative trait loci (mbQTLs) with p-value<1.0e-6 for all taxa abundance present in their analysis and kept only exposures with at least three shared mbQTLs with mean F statistics> 15. These criteria were chosen to minimize weak instrument bias and allow the use of sensitivity analysis to assess the validity of the MR assumptions. The study of Lopera-Maya et al., and Qin et al., were removed as none of the exposures satisfied our criterion (≥3 independent mbQTL at a p-value < 1e-6, with mean F-statistics > 15). In total, we included available genetic information on 48 microbial taxa abundance from the two recent GWAS studies of Kurilshikov et al., and Rühlemann et al. We then performed IVW-MR on the Wald ratio estimates for each of the 10 health outcomes (Figure 4). Out of 468 exposure-outcome tests, 32 passed a significance threshold of 0.05 and no associations were observed after Benjamini-Hochberg correction for multiple testing.

**Figure 4.**
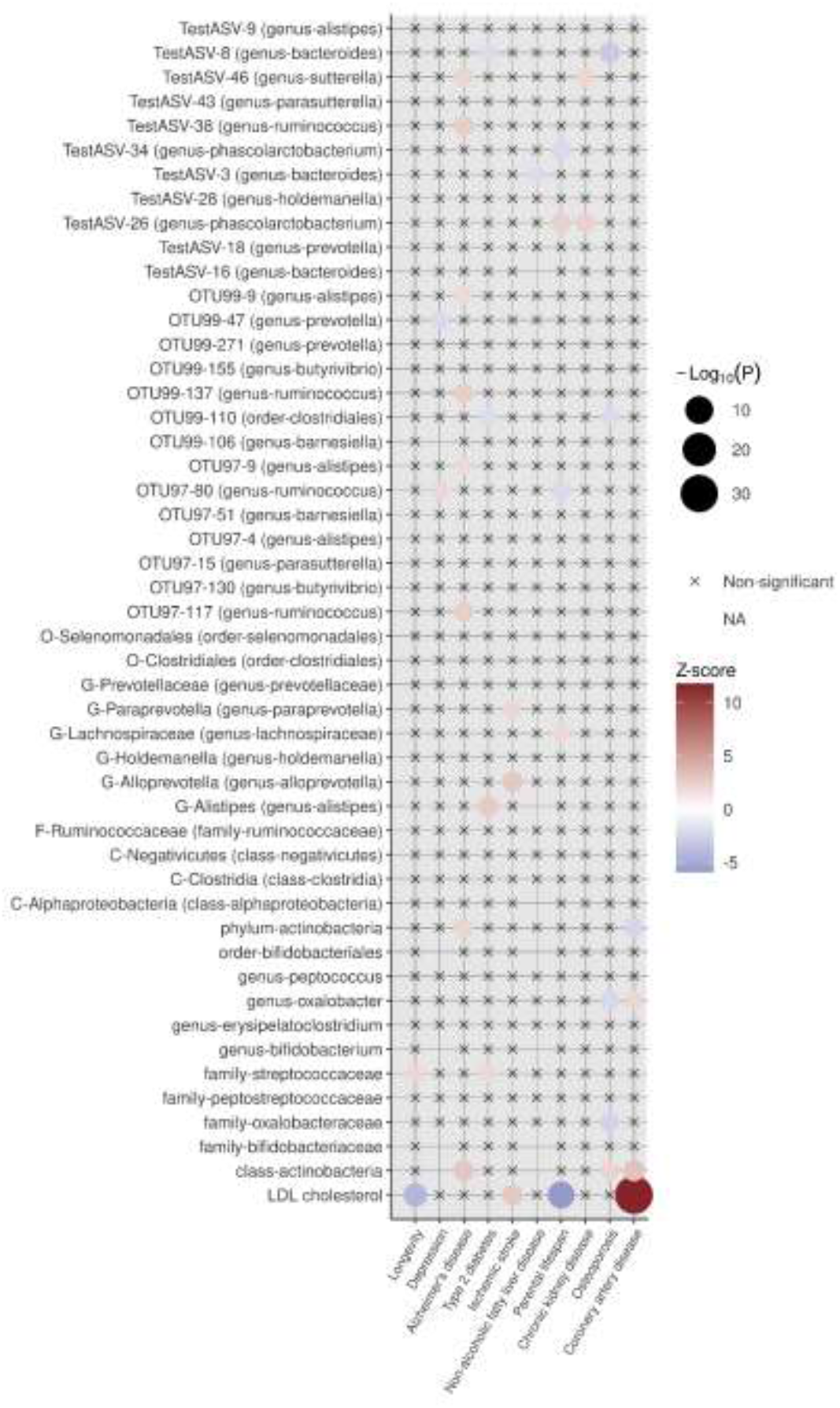
Balloon plot of the association of gut microbial taxa abundance with all 10 health outcomes. Associations with LDL cholesterol are added as positive controls. Non-available (NA) associations stem from a lack of overlapping SNPs or proxies between exposure and outcome resulting in fewer than three genetic instruments in the harmonized data set. Non-significant associations (p-value > 0.05) are depicted with crosses.

### Power calculations

All primary MR results can be retrieved in supplementary table 2. Overall, for all 582 associations tested (including IBD, microbiome associated metabolites and taxa abundance) the lowest Benjamini-Hochberg false discovery rate is 0.37 for the association between isoleucine and NALFD. Conservatively, there is no robust causal evidence for any of the 582 associations under study after considering multiple testing. Taking a more lenient approach, there are 42 associations nominally significant at p <0.05, that may be worth exploring.

In general, pleiotropy is more likely to bias estimates away from the null (Burgess et al. 2020). Therefore, a null result is generally more robust to the horizontal pleiotropy. However, concerns may arise when reporting such null findings as whether they represent a true absence of causality or are attributed to lack of power and need further exploration. To this end, we performed power analyses assessing the robustness of null findings (p>0.05) (Brion et al., 2013). We observed that our analytical framework provided enough power to detect at least moderate effect. For all 582 associations under study, we have on average 80% (±29%) power to detect a true causal effect (at beta = 0.1) (Supplementary Figure 2), meaning that an increase of 1 standard deviation unit in the exposure would change the risk of being a disease case by ∼10%. We chose this beta as a conservative estimate to what is generally observed in observational microbiome research (Kazemian et al. 2020; Zhuang et al. 2020).

### Exploration of promising findings and tests for pleiotropy

One of the key assumptions underlying MR is that genetic instruments affecting gut dysbiosis do not affect diseases or longevity by other mechanisms than the one associated with gut dysbiosis (Davies, Holmes, and Davey Smith 2018). This phenomenon is known as horizontal pleiotropy (Lawlor et al. 2019). We performed tests for pleiotropy on all potentially causal relations (primary analysis p-value <0.05) to ensure the robustness of the causal estimates. We performed robust MR analysis to estimate the robustness of our primary causal estimate to pleiotropy. We used six different methods that make different assumptions about the nature of the underlying pleiotropy: MR-Egger, the MR-Robust Adjusted Profile Score (MR-RAPS), the weighted median, the weighted mode, the MR-PRESSO and the contamination mixture approaches. Consistency across the estimates of the methods provides support to causality. Out of the 42 associations tested, eight associations had half (3/6) or more sensitivity analyses consistent with a true causal effect unlikely to be confounded by pleiotropy (p-value <0.05) (Supplementary Table 3). We therefore retained these associations for further sensitivity analysis. The association linking genetically predicted plasma isoleucine levels to NAFLD was particularly strong, where each standard deviation of genetically predicted isoleucine levels was positively associated with NAFLD (OR = 1.75 95%CI [1.18, 2.59]). The effect was consistent across most robust MR methods except MR-Egger, while the weighted mode and MR-PRESSO methods were marginally significant (Supplementary Figure 3).

### Exploration of BMI and alcohol intake as potential confounding factors

Obesity and alcohol intake frequency were recently identified as major confounding factors in the microbiome-disease associations (Vujkovic-Cvijin et al. 2020). We performed multivariable MR on the eight promising associations to determine if the genetic instruments were influenced by obesity and alcohol intake frequency. This analysis provided evidence that 4 out of 8 aforementioned reported associations might, to a certain extent, be confounded by BMI or alcohol consumption (Supplementary Figure 4). For example, the relation between NAFLD and isoleucine plasma level showed evidence of being confounded in large part by alcohol intake frequency and BMI. Overall, four associations were supported at nominal p-value. First, a 1-standard deviation (SD) increase in Actinobacteria Phylum abundance may decrease the risk of CAD OR=0.89 [0.81-0.98]) (Supplementary Figure 5), while a 1-SD increase in the *Actinobacteria* class may increase the risk of CAD (OR=1.15 [1.06, 1.26]) (Supplementary Figure 6). Second, a 1-SD increase in the abundance of the genus *Bacteroides* decreased the risk of type 2 diabetes (OR=0.93 [0.86-1.00]) (Supplementary Figure 7). Thirdly, a 1-SD increase in the genus *Phascolarctobacterium* increased lifespan (as proxied by parental lifespan) by 0.02 [0.00, 0.03] years (Supplementary Figure 8).

## Discussion

In order to determine whether previously reported studies using pre-clinical models or observational study designs in humans were consistent with causal associations, we assessed the roles of a wide range of microbial factors and eight chronic diseases and human longevity using MR. We found no association between IBD (a model of gut inflammation and dysbiosis), microbiota-associated fecal or blood metabolites and microbial abundance on these diseases and lifespan after accounting for multiple testing. Some associations, such as the relationship between Actinobacteria at Phylum or Class level and CAD and the relationship between the Bacteroides genus and T2D were nominally significant before and after sensitivity analyses and after accounting for confounders and may require further validation. Altogether, results of this study suggest that previously reported associations between the human gut microbiome and human disease might have been due to biases such as reverse causality or confounding and that the impact of gut dysbiosis on chronic diseases and human longevity may not be as prominent as previously suggested.

### Comparisons with other studies

Our results generally contrast with those from previous observational studies. First, IBD was reported in observational studies to be a risk factor of several chronic diseases including respiratory disease, arthritis, liver conditions, kidney failure, osteoporosis and depression (Bernstein et al. 2019; Xu 2018). Notably, meta-analyses of cohort studies suggest that IBD is an independent risk factor for CAD. A 2017 meta-analysis found a multivariate-adjusted independent association between IBD and CAD incident (pooled relative risk [RR]: 1.24; 95% confidence interval [CI]: 1.14 to 1.36) (W. Feng et al. 2017). The same was true for a subsequent larger meta-analysis (RR : 1.17; 95% CI[1.07, 1.27]) (Sun and Tian 2018) suggesting IBD as a potential causal factor. However, this claim is not supported by our MR analysis highlighting the potential bias induced by unmeasured confounders or participant selection bias, where participants diagnosed with IBD are more likely to be diagnosed with another disease, since they conceivably undergo clinical checkups more often.

Second, microbial metabolites have been associated with health and disease such as neurological disease, NAFLD, cardiovascular disease, survival and type 2 diabetes (Agus, Clément, and Sokol 2020; Martinez, Leone, and Chang 2017; Wilmanski et al. 2021). Notably, over 15 microbial metabolites have been identified as predictors of CAD (Q. Feng et al. 2016). Three metabolites of the dietary lipid phosphatidylcholine--choline, trimethylamine N-oxide (TMAO) and betaine--were shown to predict risk for coronary vascular disease in an independent large clinical cohort (Z. Wang et al. 2011). Moreover, increasing through dietary supplementation the level of TMAO accelerated atherosclerosis in mice (Koeth et al. 2013). By contrast, our MR analysis does not support causality between TMAO and CVD, as previously suggested in another MR investigation (Jia et al. 2019). Dietary factors could arguably act as confounding factors, since meat intake increases TMAO levels (Koeth et al. 2013).

Third, several differences in the microbial composition of diseased and healthy individuals have been identified, but causality remains to be elucidated. RCT of fecal microbiome transfer (FMT) in humans are currently employed to establish causality between microbiome and health, but few have been attempted, and even fewer have been conclusive (Depommier et al. 2019; Mocanu et al. 2021). To date, most successful randomized control study of FMT on humans has been applied to the treatment of recurrent or refractory *Clostridioides difficile* infections (Wortelboer, Nieuwdorp, and Herrema 2019) and some to ulcerative colitis (Costello et al. 2017). Mice FMTs are a valuable exploratory tool, but inference to human subjects is hazardous. Particularly, a substantial proportion of species in the human gut are not present in mice (Zheng, Baird, et al. 2017). For example, several FMT in mice from lean to obese mice resulted in improved cardiometabolic profile (Lai et al. 2018; Zoll et al. 2020), but these findings failed to replicate in humans. A systematic review of all three randomized placebo-controlled studies to treat obesity published to date found no impact of FMT on obesity, fasting plasma glucose, hepatic insulin sensitivity, or cholesterol markers across all included studies (Zhang et al. 2019). For human observational studies, multiple confounding factors could create spurious correlation between microbiome and chronic diseases, including antibiotic use, age, sex, diet, geography, BMI and alcohol intake (Kim et al. 2017). Moreover, dysbiosis could potentially be a consequence of disease states rather than a causal factor (Cani 2018).

In sum, the large proportion of “negative” findings (i.e., human gut dysbiosis may not cause chronic disease) is in line with recent literature showing an overwhelming positive publication bias in the microbiome literature (Walter et al. 2020). The publication bias can occur if studies are not adequately corrected for multiple testing and can be identified with attempt and failure to replicate (i-e winners curse bias) (Schloss 2018). For example, it was originally published that individuals with obesity were more likely to have lower bacterial diversity and relative abundances of the phylum Bacteroidetes (Turnbaugh et al. 2009), but this result failed to replicate in 9 independent cohorts (Finucane et al. 2014; Sze and Schloss 2016; Walters, Xu, and Knight 2014). Every method has its own strengths and weaknesses. Therefore, it is crucial to triangulate with different methods such as MR to address a causal research question, combining their strengths to overcome their individual weaknesses (Schloss 2018).

### Strengths and limitations

An important strength of this study is the use of the MR design with the largest publicly available GWAS datasets. Because alleles are randomly assigned and fixed at conception, biases due to confounding and reverse causality are mitigated in an MR analysis (Larsson et al. 2020). A further strength is that sample was majoritally restricted to individuals of European ancestry to reduce bias due to population stratification. However, it also restricts the generalizability of the results to this ethnic group. Lastly, power calculations were performed to ensure that results reported here were null because of a clear lack of association and not due to inadequate power to detect associations (Burgess et al. 2020).

Our study, however, has limitations. Robust genetic instruments for microbial species are challenging to find. First, microbiome heterogeneity and interindividual variability are high, substantially reducing the statistical power of microbiome GWAS analyses. Second, the phenotype is distal from individual genes making it a complex polymorphic trait with many variants of small effect size which could be prone to pleiotropy. Lastly, the twin heritability for gut microbiota taxa abundance is only on average 20% (Goodrich et al. 2014; 2016). This heritability estimate represents the upper ceiling that variance explained by genetic instruments can attain, reducing power. These factors all contributed to the fact that the number of mbQTLs identified to date is rather modest. For these reasons, we used a less stringent p-value cut-off to include a greater number of genetic variants to allow the use of sensitivity analysis and increase power. However, a less stringent p-value cut-off has the trade-off to potentially increase the chance of including false-positive effect variants which induce biases. The most important bias it introduces, the winner’s curse bias, refers to the fact that the genetic hits in discovery samples are more likely to be false positive, adding noise to the analysis which will typically bias MR results toward the null. Second in importance, the weak instrument bias occurs when the variance explained by the instrument and the sample size are low (Burgess, Thompson, and CRP CHD Genetics Collaboration 2011). In the two sample MR setting, it will bias towards the null (Burgess, Davies, and Thompson 2016). Third in importance, invalid instruments are pleiotropic variants that affect the outcome via another pathway than the one going through the exposure. Pleiotropic variants can invalidate the MR estimate and usually bias away from the null (Slob and Burgess 2020). We minimized weak instrument biases by including only exposures with genetic instruments with an average F-statistic above 15. We minimized the propensity of the results to be biased by pleiotropic variants by including only exposures with three genetic instruments minimum and performed sensitivity analysis. A second limitation is that the microbiome GWAS included in the current analysis did not target the entire 16S gene, which greatly diminished their ability to achieve a sufficient taxonomic resolution to identify potential therapeutic targets. The meta-analysis by Ruhlemann targeted the V1-V2 subregion while the meta-analysis by Kurilshikov et al. included mostly the V4 subregion and to a lesser extent the V1-V3 subregion. Targeting only 16S subregions such as V4 leads to lower taxonomic resolution achieved compared to sequencing the full V1-V9 16S gene (Johnson et al. 2019). Indeed, using a variable region as a surrogate for the entire 16S gene only allows for the identification of taxa at the genus level or above (Johnson et al. 2019). Being confident at the genus level provides little information to shape disease treatment. Indeed, within a high-level taxon such as a phylum, some species may have a positive correlation with a disease, but some neutral or negative. For example, in our study, the phylum Actinobacteria was potentially protective for CAD, while the subsequent level, the class Actinobacteria was a potential risk factor for the same disease. Employing third-generation technologies has the potential to allow the sequencing of the full 16S gene in a high throughput manner, and improve taxonomic discrimination.

## Conclusion*s*

Using MR, an approach less subject to reverse causality and confounding factors in comparison to traditional methods, we showed that several features of human gut dysbiosis including IBD, plasma and fecal metabolites as well as a large proportion of associations between 48 microbial taxa and 8 chronic diseases and longevity were not significant. While further study is needed, these results do not support causal impact of microbiome and microbial metabolites on human chronic diseases and longevity. As the microbiome field matures, the use of more populous microbiome GWAS study taking advantage of discriminatory potential of the full 16S gene is warranted.

## Methods

### Study exposures

We derived our 60 exposures of interest from six different publicly available data sources (supplementary table 4). Genetic instruments for IBD were obtained from a meta-analysis of 38,155 cases and 48,485 controls. Data originated from a meta-analysis of genome-wide association from seven Crohn’s disease and eight ulcerative colitis GWAS collections with genome-wide genotyping or immunochip data from individuals of European descent from 15 countries in Europe, North America and Oceania (Liu et al., 2015a). Diagnosis of IBD was based on radiological, endoscopic or histopathological evaluation. We included independent genetic variants (r2 ≤ 0.001) strongly associated with IBD (p-value <5e-8).

Genetic instruments for fecal propionate and PWY-5022 were obtained from a GWAS on 952 normoglycemic participants of the LifeLines-DEEP cohort, a population-based cohort from northern Netherlands (age ranges 18–84 years) (Sanna et al. 2019). Fecal propionate levels were measured by gas chromatography-mass spectrometry (GCMS). The functional pathway PWY-5022 was obtained with HUMAnN2 (v 0.4.0) (Franzosa et al., 2018) and MetaCyc metabolic-pathway database (Vatanen et al. 2016). Genotyping was carried out with two Illumina arrays, HumanCytoSNP-12 BeadChip and ImmunoChip.

Genetic instruments for plasma TMAO, carnitine, betaine, choline and indole-3-propionate were extracted from a GWAS conducted in 2076 participants from European ancestry from the Framingham Heart Study (FHS) offspring cohort (Rhee et al. 2013). The FHS offspring cohort is a prospective community-based cohort from Framingham, Massachusetts, USA. Children of the spouse of the FHS study were recruited in 1971. Metabolites profiling was performed by liquid chromatography-mass spectrometry (T. J. Wang et al. 2011). Genotyping was conducted using the Affymetrix 500K mapping array and the Affymetrix 50K gene-focused MIP array. The participants all provided their informed consent and the study was approved by the Boston University Medical Center.

Genetic instruments for plasma branched-chain amino acids (leucine, isoleucine and valine) and acetate were extracted from a meta-analysis of GWAS conducted on 10 European cohorts totalizing 24,925 individuals (Kettunen et al. 2016). Human blood metabolites were quantified with quantitative high-throughput NMR metabolomics platform.

Genetic instruments for bacterial taxon were extracted from a GWAS of bacterial taxon abundances of 8,956 German individuals from the PopGen (population-based cohort), the FoCus (population registry based), the KORA FF4 (population-based adult cohort initiated in 1984) and the SHIP cohort (longitudinal population-based cohort) (Rühlemann et al. 2021). Human Genotyping and fecal microbial 16S rRNA gene surveys were performed using multiple arrays. Additionally, other genetic instruments for bacterial taxon were extracted from a meta-analysis conducted by the MiBioGen consortium on 16S fecal microbiome data from 18,340 individuals (24 cohorts) (Kurilshikov et al. 2021). All cohorts implemented the standardized 16S processing pipeline that uses SILVA as a reference database, with truncation of the taxonomic resolution of the database to genus level. Cohorts were Middle Eastern, East Asian, American Hispanic/Latin, African American and admixed, although the majority of the sample (more than 72%) came from European descent. Because the effect sizes of mbQTLs were not available in the summary data, we estimated beta from the *z* statistic using the following equation. *b = z* * *SE* where *b* is the beta, *z* is the z-statistics and *SE* is the standard error. We estimated *SE* with the following equation 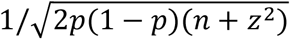 where *p* is minor allele frequency and *n* is sample size. Allele frequency was also not available in the GWAS summary statistics. It was estimated from the reference panel used by 23 out of 24 cohorts, the Haplotype Reference Consortium (HRC 1.1 reference panel) (McCarthy et al. 2016).

### Study Outcomes

We used publicly available GWAS summary statistics from the largest studies of chronic disease that have been previously linked to the human gut microbiota (with the exception of NAFLD which was performed for the purpose of this study) (Supplementary Table 5). **CAD.** GWAS summary statistics for CAD were obtained from a GWAS on 88,192 cases and 162,544 controls from CARDIoGRAMplusC4D and UK Biobank (van der Harst and Verweij 2018). **Ischemic stroke.** GWAS summary statistics for ischemic stroke were obtained in 67,162 cases/454,450 controls from the MEGASTROKE consortium (Malik et al. 2018). **Alzheimer’s disease.** GWAS summary statistics for Alzheimer’s Disease were obtained from a meta-analysis of the PGC-ALZ, IGAP, ADSP consortium and UK Biobank comprising 71,880 cases/383,378 controls (Jansen et al. 2019). **Depression.** GWAS summary statistics were obtained for depression from a meta analysis of PGC/UKB (excluding 23andme) including 500,199 (170,756 cases and 329,443 controls) individuals (Howard et al. 2019). **Osteoporosis.** Genetic association estimates for osteoporosis were obtained in the UK Biobank (PheCode_743.1) comprising of 7547 cases and 455,386 controls (Zhou et al. 2018). **Type 2 diabetes.** GWAS summary statistics for type 2 diabetes were obtained from the DIAGRAM consortium (74,124 cases/824,006 controls) (Mahajan et al. 2018). **Chronic kidney disease.** GWAS summary statistics for chronic kidney disease were obtained from the CKDGen consortium including 41,395 cases and 439,303 controls (Wuttke et al. 2019). **Parental lifespan and longevity.** We also used publicly available GWAS summary statistics for human longevity-related outcomes namely parental survival (from a meta-analysis of the UK Biobank and the LifeGen consortium of 26 population cohorts [n=1,012,240; all of European ancestry]) (Timmers et al. 2019) and human longevity from a meta-analysis of 18 European cohorts (Deelen et al. 2019). Longevity was defined as individuals surviving at or beyond the age corresponding to the 90th survival percentile and controls whose age at death or at last contact was at or below the age corresponding to the 60th survival percentile (11,262 cases/25,483 controls). **Non-alcoholic fatty liver disease.** To obtain a comprehensive set of NAFLD GWAS summary statistics, we performed a GWAS meta-analysis of four cohorts as previously described (Ghodsian et al. 2021). Data sources included the Electronic Medical Records and Genomics (eMERGE) network, the UK Biobank, the Estonian Biobank and FinnGen. The NAFLD GWAS (1106 cases and 8571 controls) in the eMERGE network has previously been published and summary statistics were made available (Namjou et al. 2019). We performed a new GWAS for NAFLD in the UK Biobank that included 2558 cases and 395,241 controls (data application number 25205). UK Biobank has approval from the North West Multi-Centre Research Ethics Committee. We also performed a GWAS for NAFLD using SAIGE in the Estonian Biobank (4119 cases and 190,120 controls. This was approved by the Research Ethics Committee of the University of Tartu (Approval number 288/M-18). Finally, we used publicly available NAFLD GWAS summary statistics of the FinnGen cohort (651 cases and 176,248 controls). The total study sample for the GWAS meta-analysis included 8434 cases and 770,180 controls. This project was approved by the Institutional Review Board of the Quebec Heart and Lung Institute.

### Selection of Genetic Variants and Variants Harmonization

We first identified all SNPs associated with exposures. Summary parameters for genetic instrument selection can be found at supplementary table 6. These SNPs were then clumped using the 1,000-Genome Project Phase 3 European LD reference panel to make sure instrumental variables were independent (Auton et al., 2015) with a 10 Mb window and pairwise linkage disequilibrium (LD) r2 <0.01. This step was implemented with the *TwoSampleMR* package in R (Hemani et al. 2018). Because of their known association with pleiotropic pathways, we excluded from the analysis all SNPs of the *HLA, ABO* and *APOE* genetic regions. We then performed Steiger filtering to remove variants with evidence of a stronger association with the outcome than its association with the exposure (Hemani, Tilling, and Davey Smith 2017). Instrument strength was quantified using the F-statistic (Burgess, Thompson, and CRP CHD Genetics Collaboration 2011), and the variance explained was quantified using the R^2^ (Pierce, Ahsan, and VanderWeele 2011).

Variant harmonization was performed by aligning the betas of different studies on the same effect allele with the *TwoSampleMR* package (Hemani et al. 2018). We inferred positive strand alleles, using allele frequencies for palindromes whenever possible. This method is effective when effect allele frequency difference is large, but may not, however, be a reliable indicator of reference strands when it is close to 0.5 (Hartwig et al. 2016). Consequently, palindromic SNPs with MAF > 0.3 were removed from the analysis. When a particular exposure SNP was not present in the outcome dataset, we used proxy SNPs instead (r2 > 0.8). We obtained the LD matrix of the 1000 genomes project - European sample of the Utah residents from North and West Europe via the LDlink API (Machiela and Chanock 2015). We interrogated the LDlink API with the *LDlinkR* package (Myers, Chanock, and Machiela 2020). We kept only the results based on at least three independent shared SNPs with mean F statistics> 15 to reduce weak instrument bias and allow for sensitivity analysis.

### Statistical Analyses

Genetic correlation was performed using linkage disequilibrium score regression (LDSC). LDSC computes the genetic covariance between traits based on genome wide summary statistics obtained from GWAS (Bulik-Sullivan et al. 2015). The complete summary statistics of IBD and all outcomes were used. The LDScore regression function was fit using the GenomicSEM R package (Grotzinger et al. 2019) except for osteoporosis, where the genetic correlation was computed with the LD Hub database (Zheng, Erzurumluoglu, et al. 2017). LD Hub includes GWAS publicly available summary statistics on hundreds of human traits and enables the assessment of LD score regression among those traits.

We conducted primary MR analysis on each outcome and exposure association. As primary method for causal inference, we performed the IVW method with multiplicative random effects with a standard error correction for under dispersion as recommended by recent MR guidelines (Burgess et al. 2020). The IVW-MR combines the ratio estimates from each genetic instrument in a meta-analysis model by giving more weight to the ratio estimates with lower variance (Burgess, Small, and Thompson 2017). A total of 582 primary analyses were performed: 60 exposures * 10 outcomes—18 exposures/outcomes with fewer than three overlapping genetic instruments. We applied a Benjamini Hochberg correction for multiple testing using a false discovery rate of 5% to reduce the propensity of false positive finding. All continuous exposure estimates are normalized and reported on a standard deviation scale. For dichotomous traits (all outcomes except longevity and IBD as exposure), we transformed ORs and CIs to effect sizes and standard error when it was not already done. Longevity estimates were reported by additional year of lifespan.

For nominally significant associations (IVW-MR p-value <0.05) sensitivity analyses to estimate the robustness of our primary causal estimate were performed. We used 6 different robust methods that make different assumptions about the nature of the underlying pleiotropy. As a general test to the presence of pleiotropy, we used Egger intercept regression (MR-Egger), the MR-Robust Adjusted Profile Score (MR-RAPS) approaches and the contamination mixture approach. MR-Egger is similar to the IVW method except the regression model estimates an intercept (Bowden et al., 2015). An intercept significantly different from zero gives indication of pleiotropy (Burgess & Thompson, 2017). The MR-Egger method gives consistent estimates of the causal effect under the InSIDE assumption, which states that pleiotropic effects of genetic variants must be uncorrelated with genetic variants—exposure association (Slob and Burgess 2020). MR-RAPS models the pleiotropic effects of genetic variants directly using a linear random effects distribution. This method provides an unbiased estimate when there is no pleiotropy (Zhao et al. 2018). The contamination mixture provides consistent estimate under the plurality valid assumption (Slob and Burgess 2020). It assumes that not all genetic variants are valid IVs and runs a likelihood function to categorize genetic instruments as valid or invalid. As a general test of robustness of the IVW-MR estimates, we used the weighted median and the weighted mode methods. The weighted median allows an unbiased estimate to be obtained if the “majority valid” assumption is upheld, that is if up to 50% of the weight comes from variants that are valid IVs (Bowden et al. 2016). The weighted mode estimates the causal effect from the value taken by the largest number of genetic variants. This method allows the true causal estimates to be unbiasedly estimated if the “plurality valid” assumption is upheld, that is only 40% of the genetic variants are valid instruments and no larger group with the same estimate exists (Hartwig, Davey Smith, and Bowden 2017). Finally, as a general test to the presence of outliers, we used the outlier-robust method MR-PRESSO, which is a simulation approach where genetic variants are removed based on their contributions to heterogeneity (Verbanck et al. 2018). This method provides consistent estimates under the same assumptions as the IVW-MR method for the set of genetic variants that are not identified as outliers (Slob and Burgess 2020). We also provide a measure of the heterogeneity between variant-specific causal estimates, with Cochran’s Q statistic (Bowden et al. 2017).

Obesity and alcohol intake frequency were recently identified as major confounding factors in the gut-disease associations (Vujkovic-Cvijin et al. 2020) because they are both to some extent associated with the health outcome under study (Blüher 2019; Jani et al. 2021) while potentially simultaneously influencing microbiome composition (Dubinkina et al. 2017). In observational studies, adding BMI and alcohol intake frequency are the most important predictors of microbiota composition and health and adding them as covariates in linear mixed-effect models reduced the numbers of spurious microbiome health associations that are identified as significant (Vujkovic et al. 2021). To account for this, we performed multivariable MR as a sensitivity analysis to correct for measured confounders (Gormley et al. 2020). BMI and alcohol intake frequency GWAS were obtained from accessing the MR-BASE database from the *TwoSampleMR* package (Walker et al. 2019). Both GWAS were derived from the UK biobank. Multivariate MR IVW estimates were computed using the *MendelianRandomization* package (Yavorska and Burgess 2017).

### Institutional Review Board Approval

All GWAS summary statistics that were used we in the public domain with the exception of the NAFLD GWAS meta-analysis that is based on two new analysis from the UK Biobank and the Estonian Biobank. The UK Biobank was performed using data application number 25205. UK Biobank has approval from the North West Multi-Centre Research Ethics Committee. This analysis of the Estonian Biobank was approved by the Research Ethics Committee of the University of Tartu (approval number 288/M-18). For the other publicly available GWAS summary statistics, all participants provided informed consent and study protocols were approved by their respective local ethical committee (Deelen et al. 2019; Howard et al. 2019; Jansen et al. 2019; Kettunen et al. 2016; Kurilshikov et al. 2021; J. Z. Liu et al. 2015b; Mahajan et al. 2018; Rhee et al. 2013; Rühlemann et al. 2021; Sanna et al. 2019; Timmers et al. 2019; van der Harst and Verweij 2018; Wuttke et al. 2019). This project was approved by the Institutional Review Board of the Quebec Heart and Lung Institute.

## Data Availability

All data used in this study are in the public domain. Supplementary tables 4 and 5 describe the data used and relevant information to retrieve the summary statistics.

## Data Availability

All data used in this study are in the public domain (or will be shortly). Supplementary tables 4 and 5 describe the data used and relevant information to retrieve the summary statistics.

## Code Availability

Code was performed in the R V.4.0.0 computing environment using publicly accessible functions from the *TwoSampleMR* V.0.5.5 https://github.com/MRCIEU/TwoSampleMR, the *MendelianRandomization V.0.5.1* https://cran.rproject.org/web/packages/MendelianRandomization/index.html and the *data.table V.1.14.0* https://github.com/Rdatatable/data.table *packages. The tidyverse V.1.3.1* collection of R packages was also used (Wickham et al. 2019).

## Acknowledgements

We would like to thank all study participants as well as all investigators of the studies that were used throughout the course of this investigation. EG holds a masters research award from the *Fonds de recherche du Québec: Santé*. (FRQS). BJA and ST hold junior scholar awards from the FRQS. Part of this study was supported by the European Union through the European Regional Development fund. The work of Estonian Genome Center, Univ. of Tartu has been supported by the European Regional Development Fund and grants No. GENTRANSMED (2014-2020.4.01.15-0012), MOBERA5 (Norface Network project no 462.16.107) and 2014-2020.4.01.16-0125. This study was also funded by the European Union through Horizon 2020 research and innovation programme under grant no 810645 and through the European Regional Development Fund project no. MOBEC008 and Estonian Research Council Grant PUT1660.

