## Supplementary Figures for "A multidimensional Mendelian randomization study on the impact of gut dysbiosis on chronic diseases and human longevity"

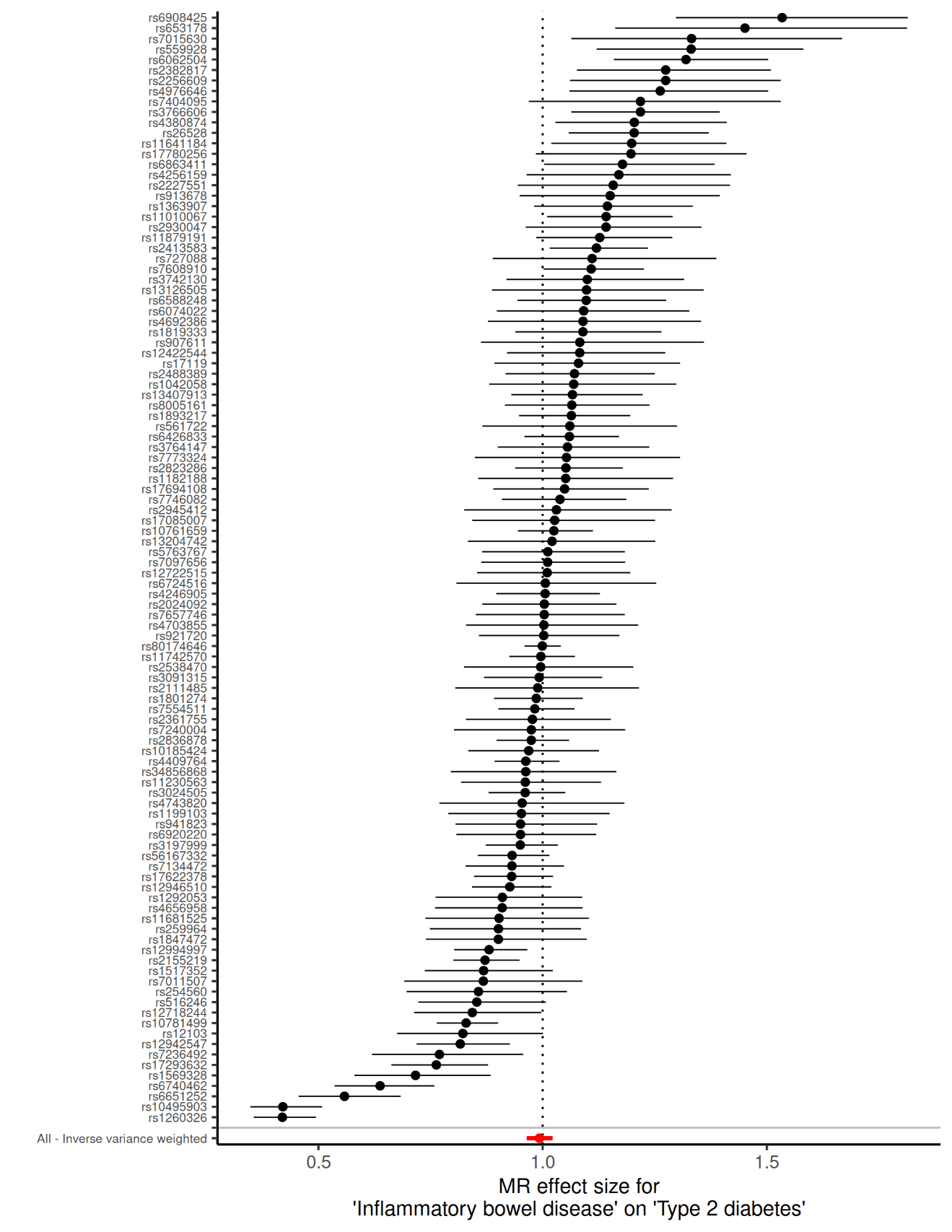

**Supplementary Figure 1A. Wald ratio of each individual SNP and their inverse variance weighted meta-analysis for the effect of inflammatory bowel disease on type 2 diabetes.**

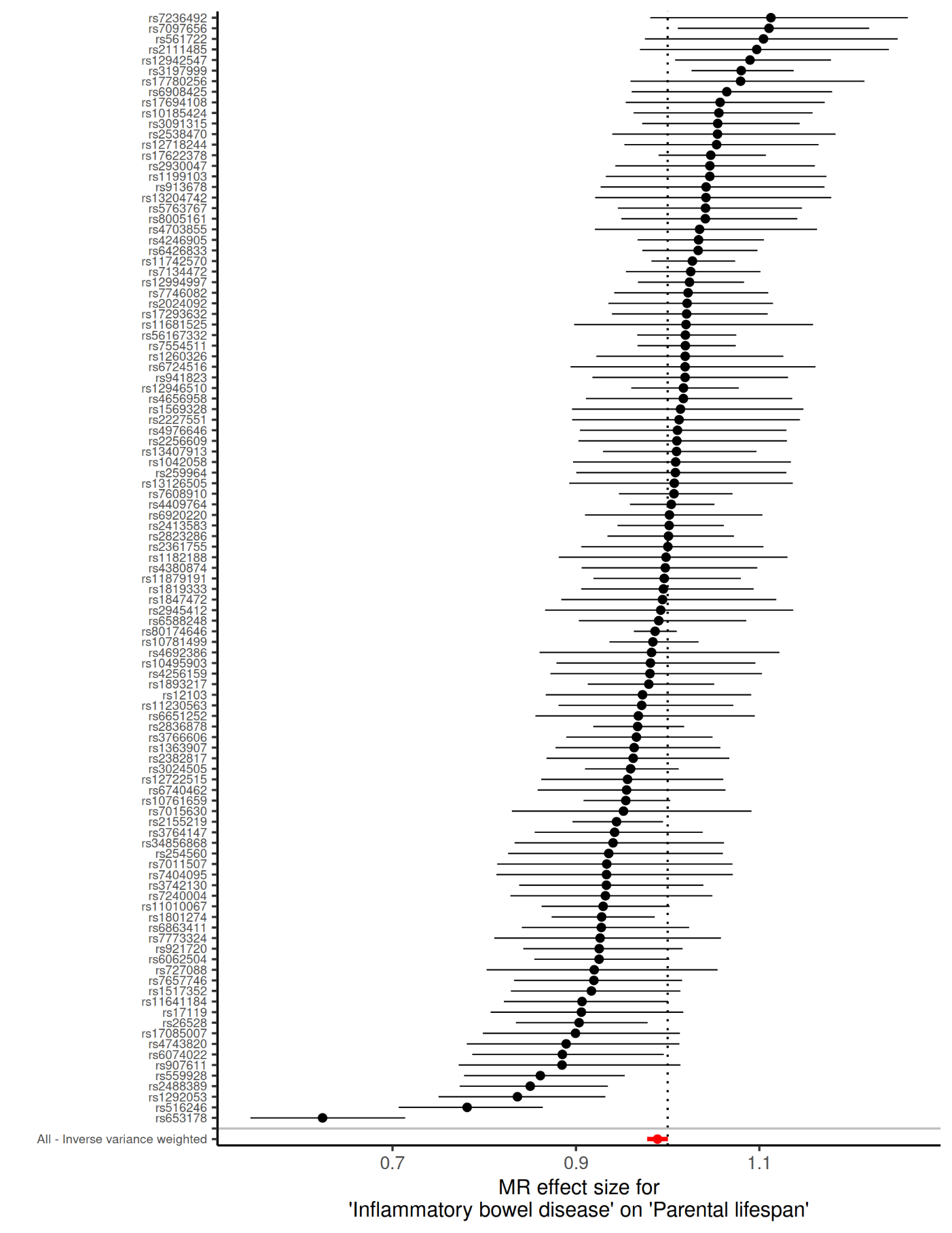
 **Supplementary Figure 1B. Wald ratio of each individual SNP and their inverse variance weighted meta-analysis for the effect of inflammatory bowel disease on parental lifespan**

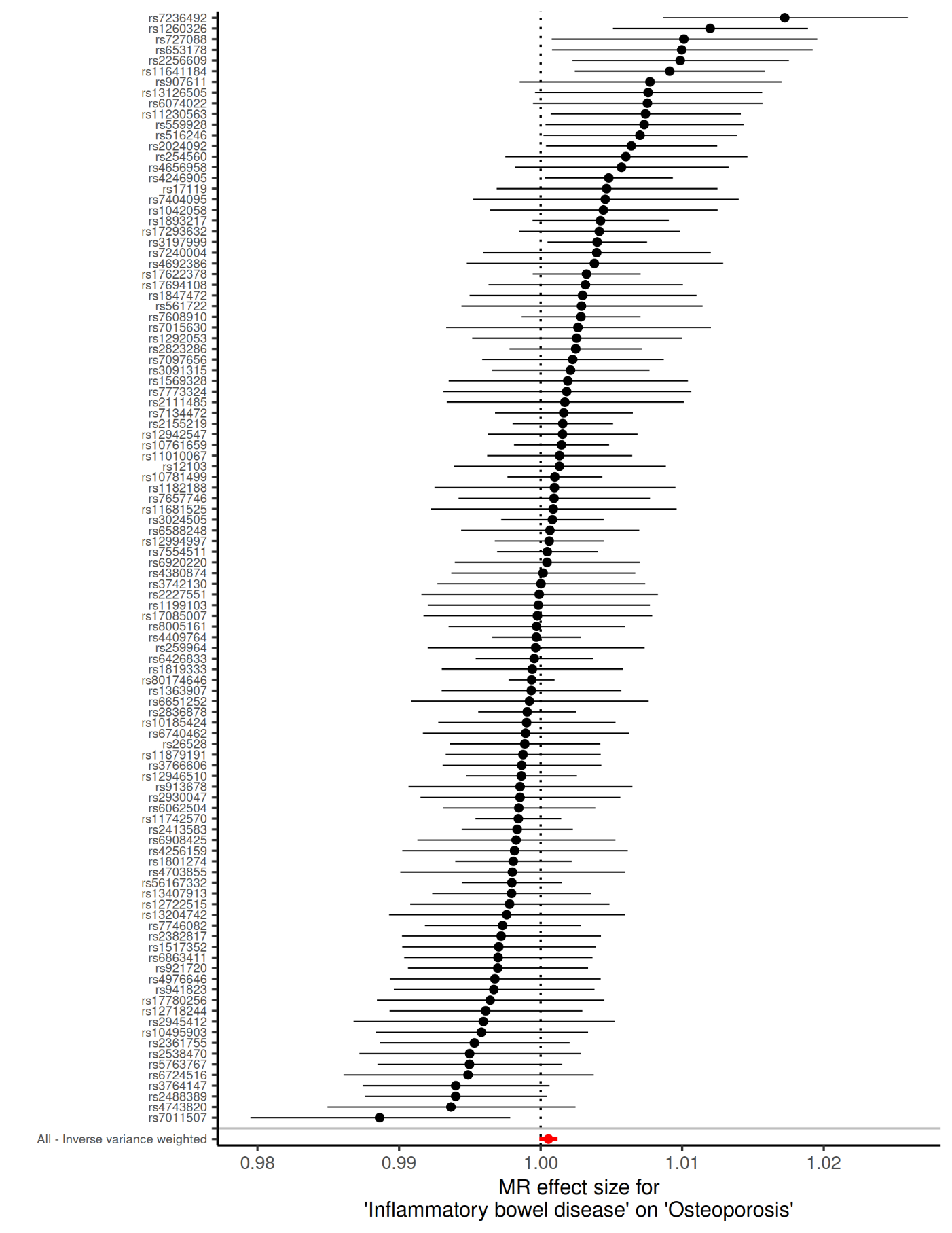
 **Supplementary Figure 1C. Wald ratio of each individual SNP and their inverse variance weighted meta-analysis for the effect of inflammatory bowel disease on osteoporosis**

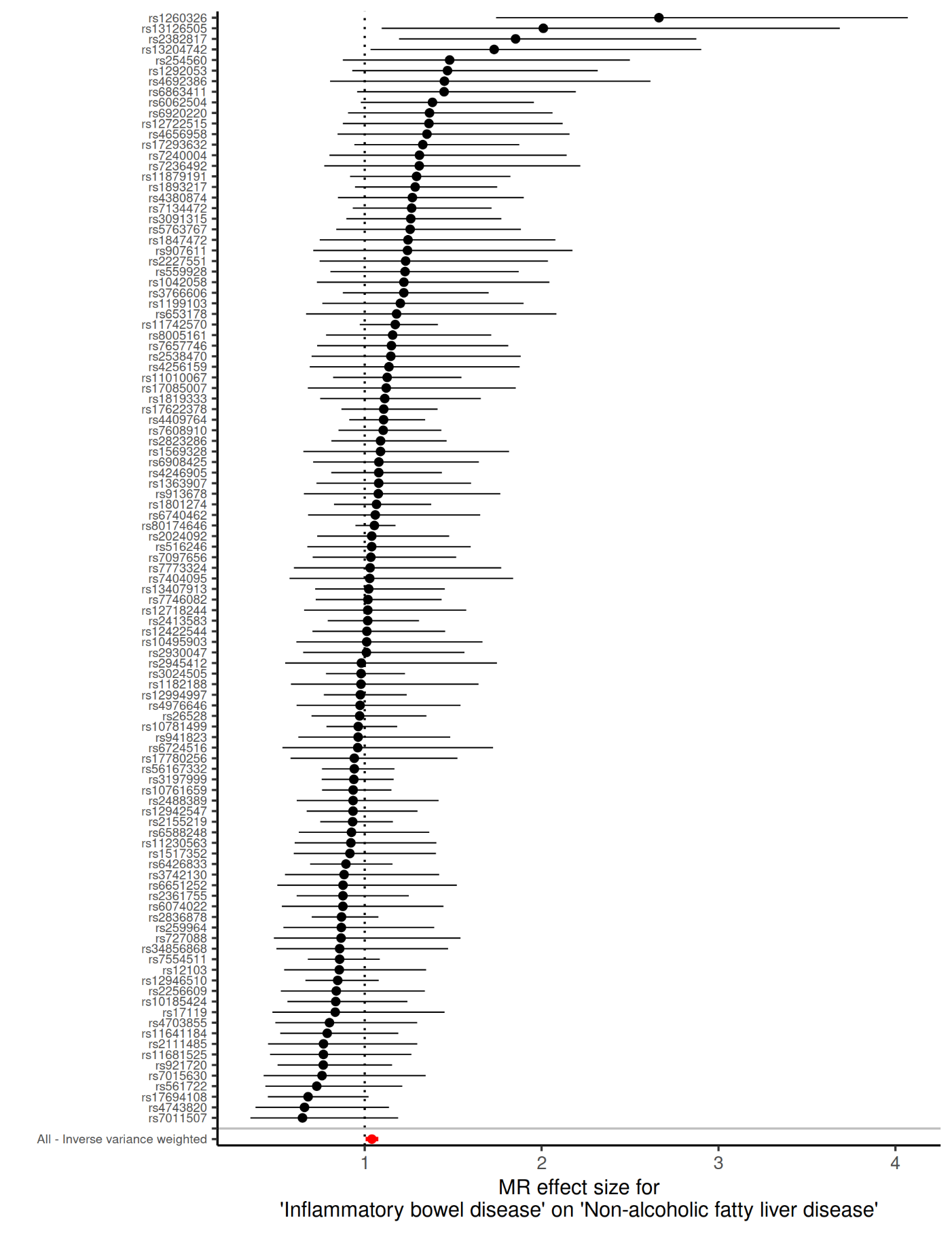
 **Supplementary Figure 1D. Wald ratio of each individual SNP and their inverse variance weighted meta-analysis for the effect of inflammatory bowel disease on non-alcoholic fatty liver disease**

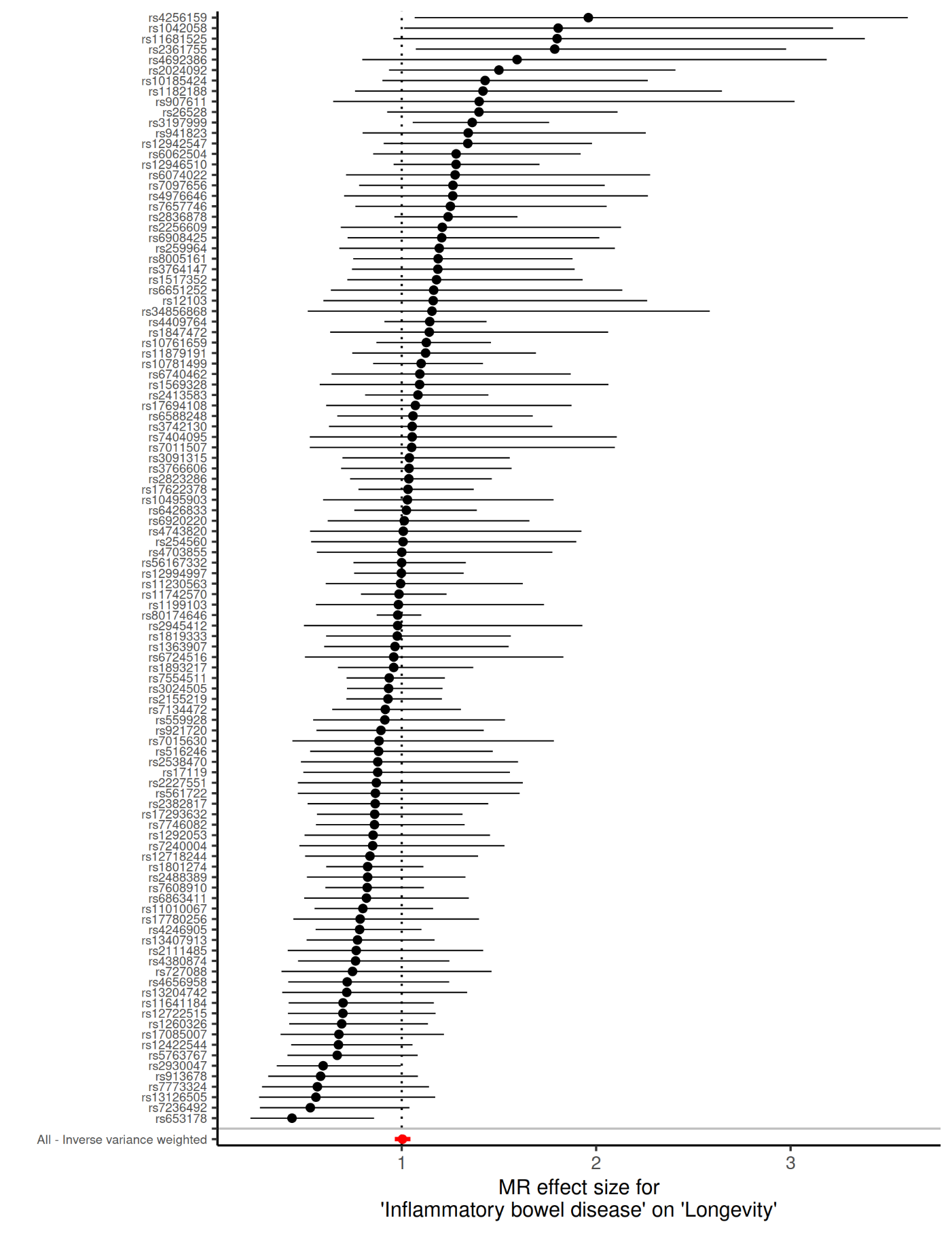
 **Supplementary Figure 1E. Wald ratio of each individual SNP and their inverse variance weighted meta-analysis for the effect of inflammatory bowel disease on longevity.**

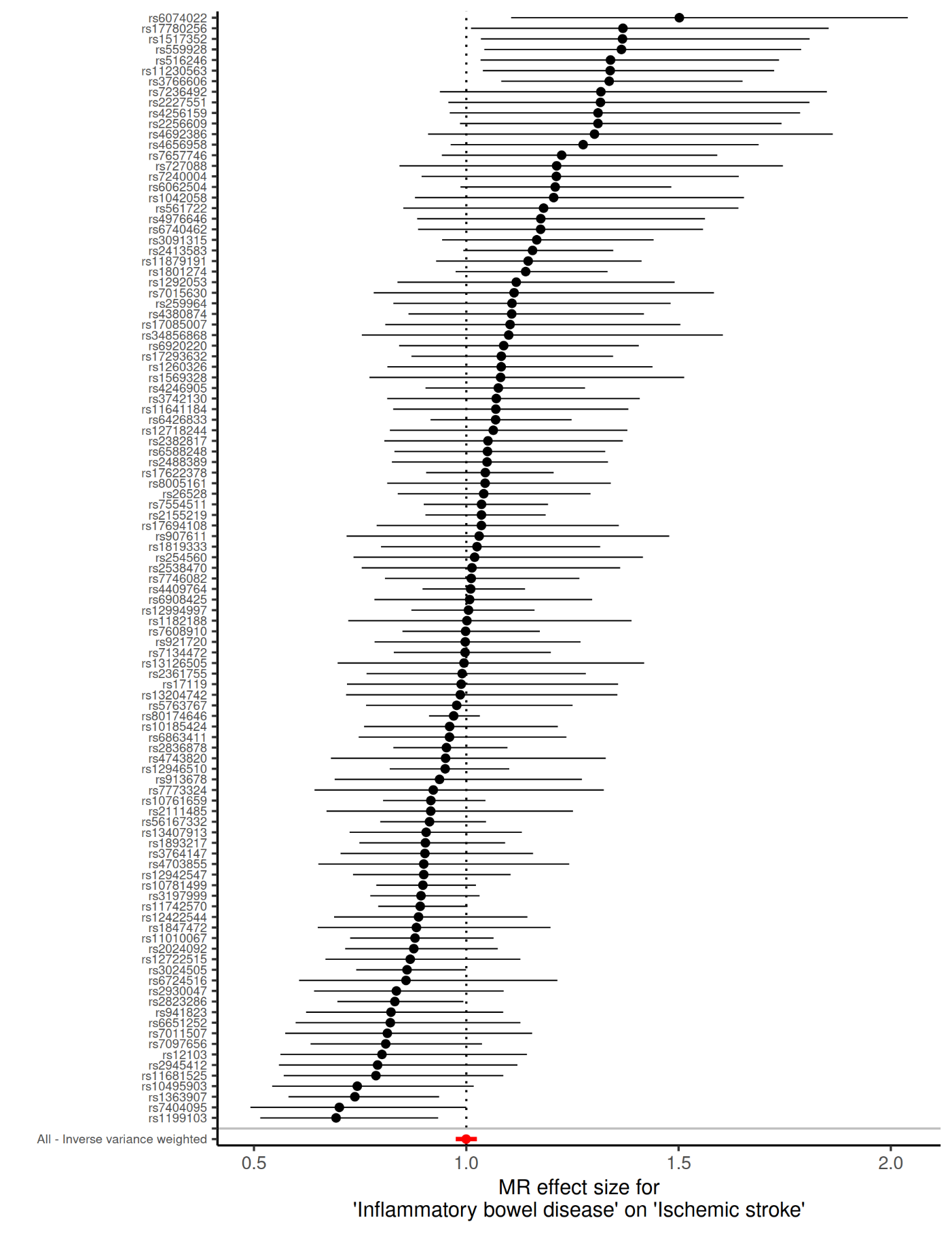
 **Supplementary Figure 1F. Wald ratio of each individual SNP and their inverse variance weighted meta-analysis for the effect of inflammatory bowel disease on ischemic stroke.**

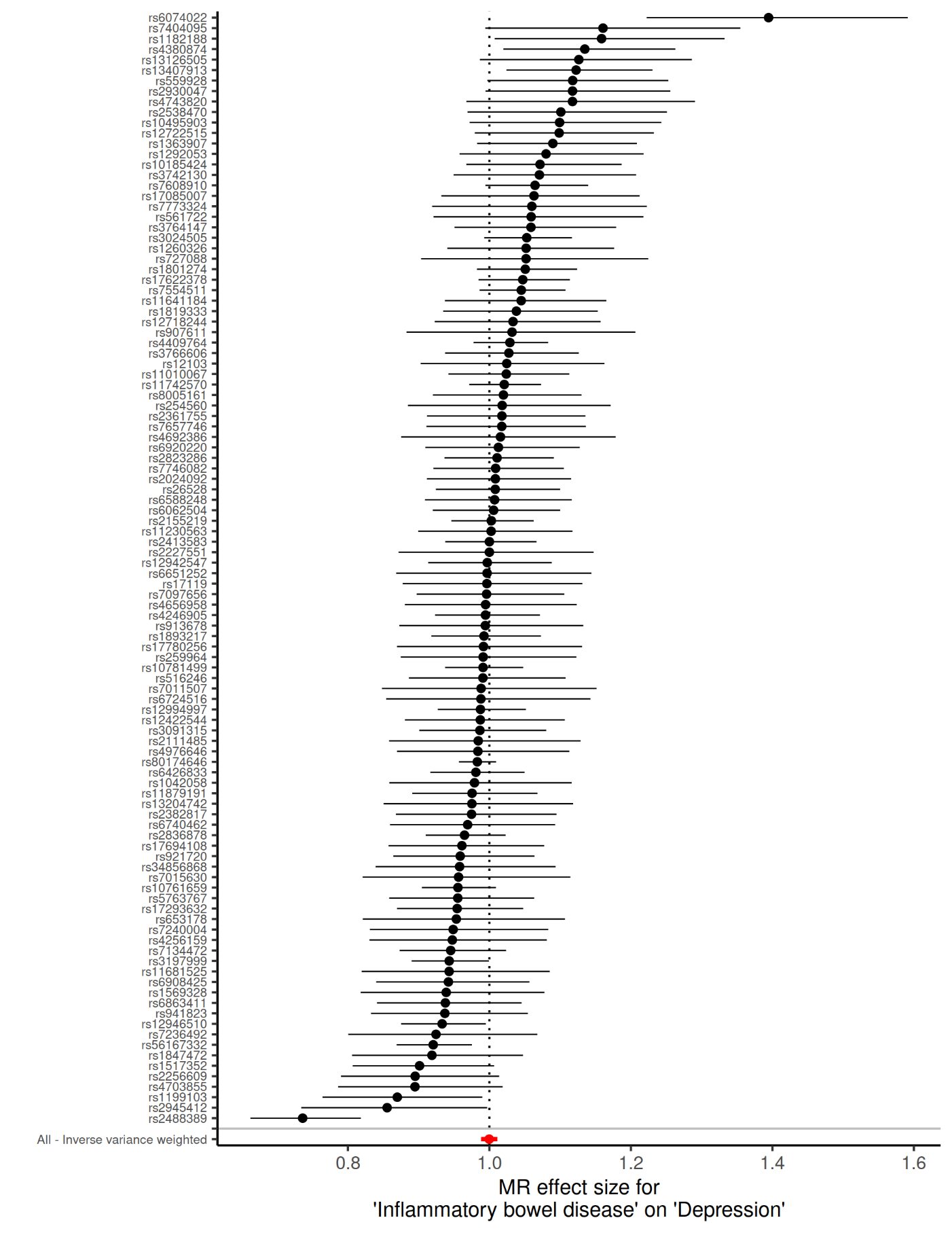
 **Supplementary Figure 1G. Wald ratio of each individual SNP and their inverse variance weighted meta-analysis for the effect of inflammatory bowel disease on depression.**

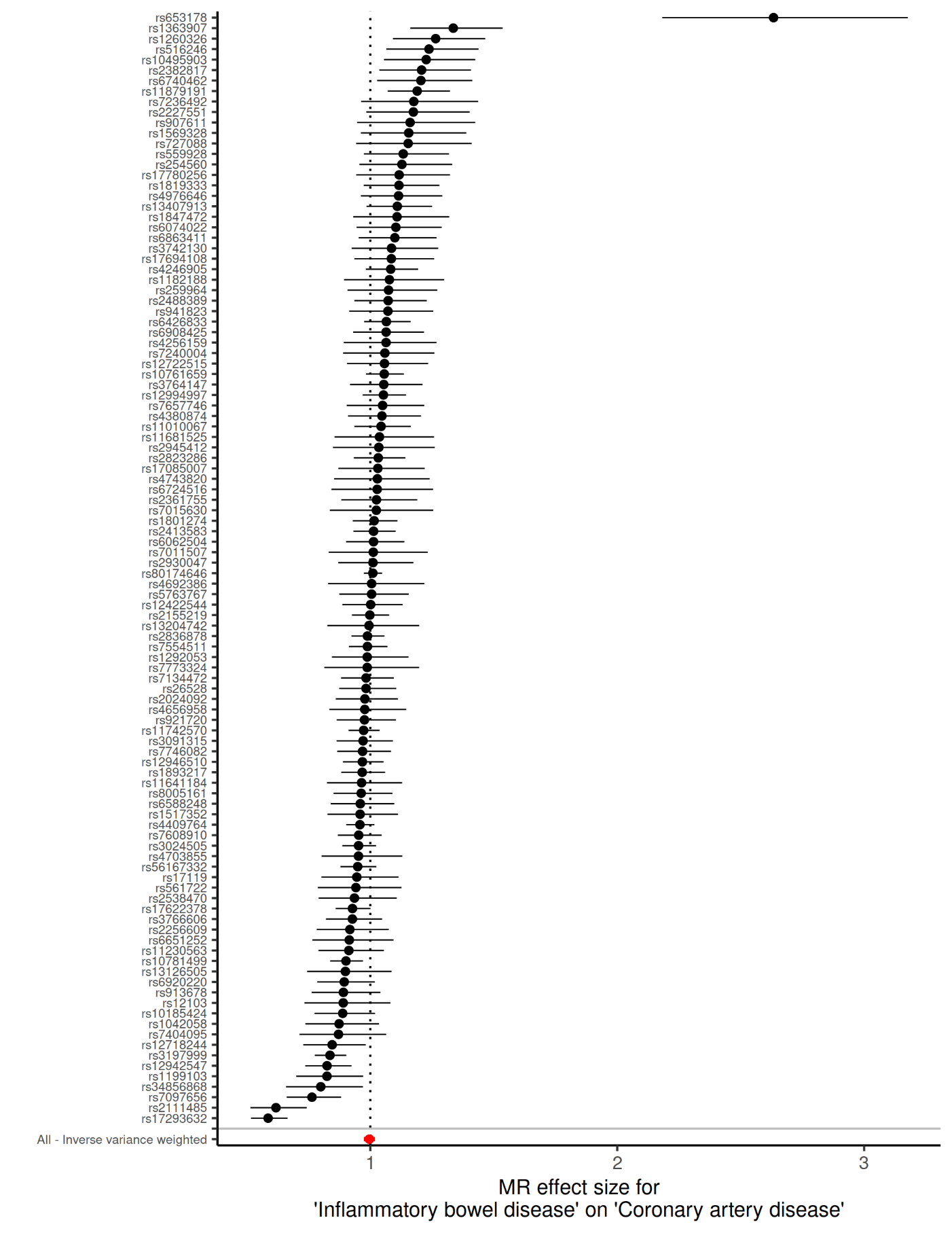
 **Supplementary Figure 1H. Wald ratio of each individual SNP and their inverse variance weighted meta-analysis for the effect of inflammatory bowel disease on coronary artery disease**

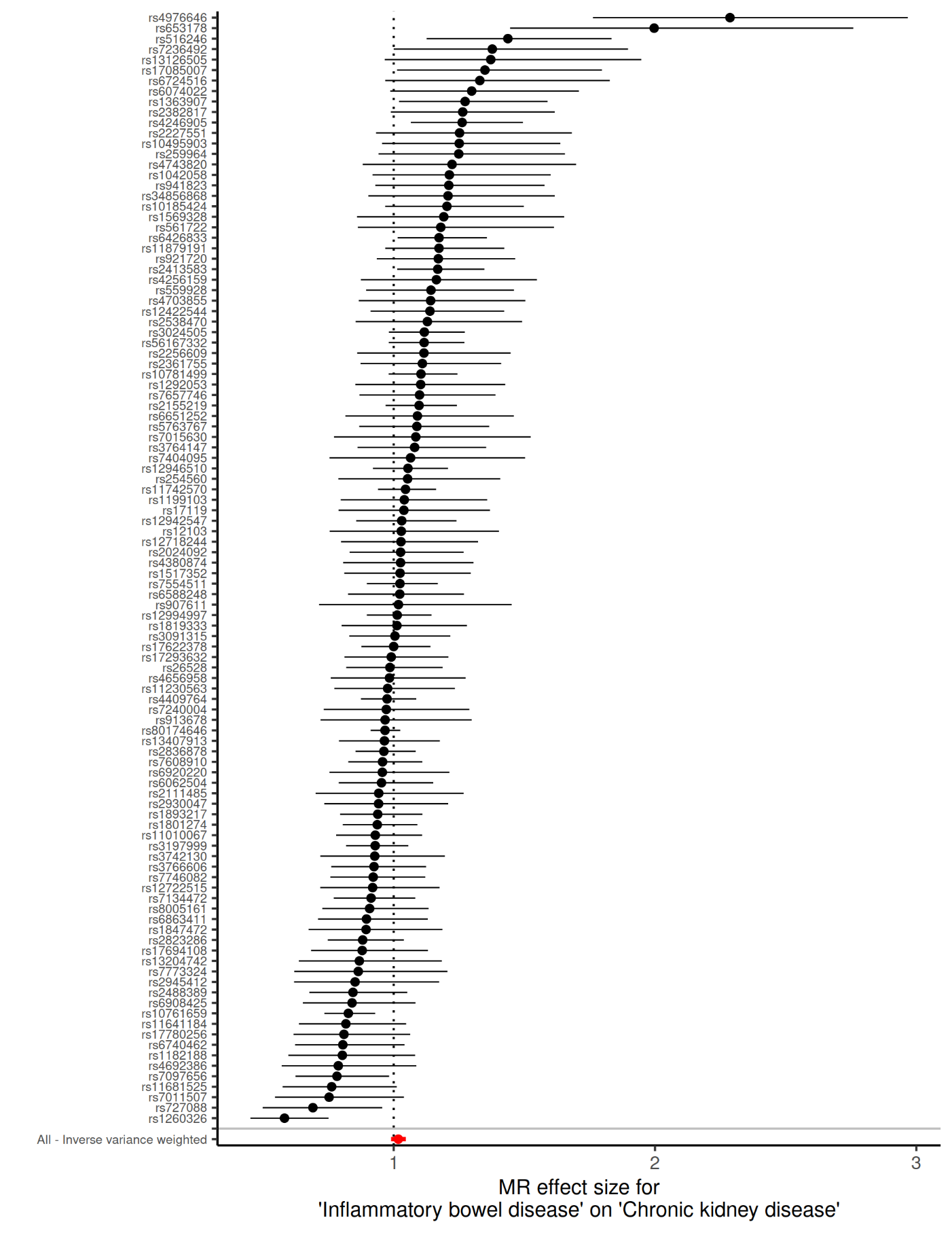
 **Supplementary Figure 1I. Wald ratio of each individual SNP and their inverse variance weighted meta-analysis for the effect of inflammatory bowel disease on chronic kidney disease**

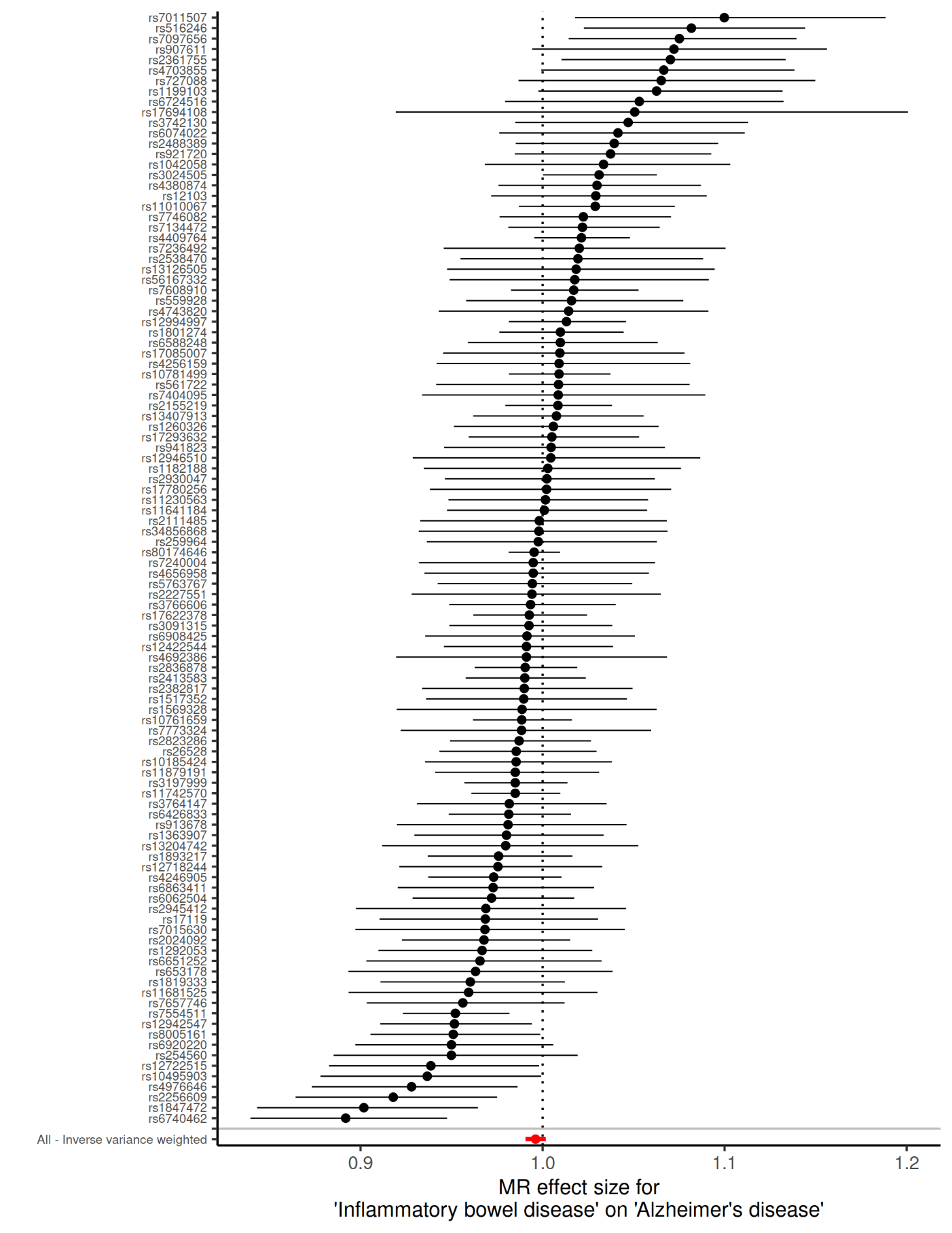
 **Supplementary Figure 1J. Wald ratio of each individual SNP and their inverse variance weighted meta-analysis for the effect of inflammatory bowel disease on Alzheimer’s disease**

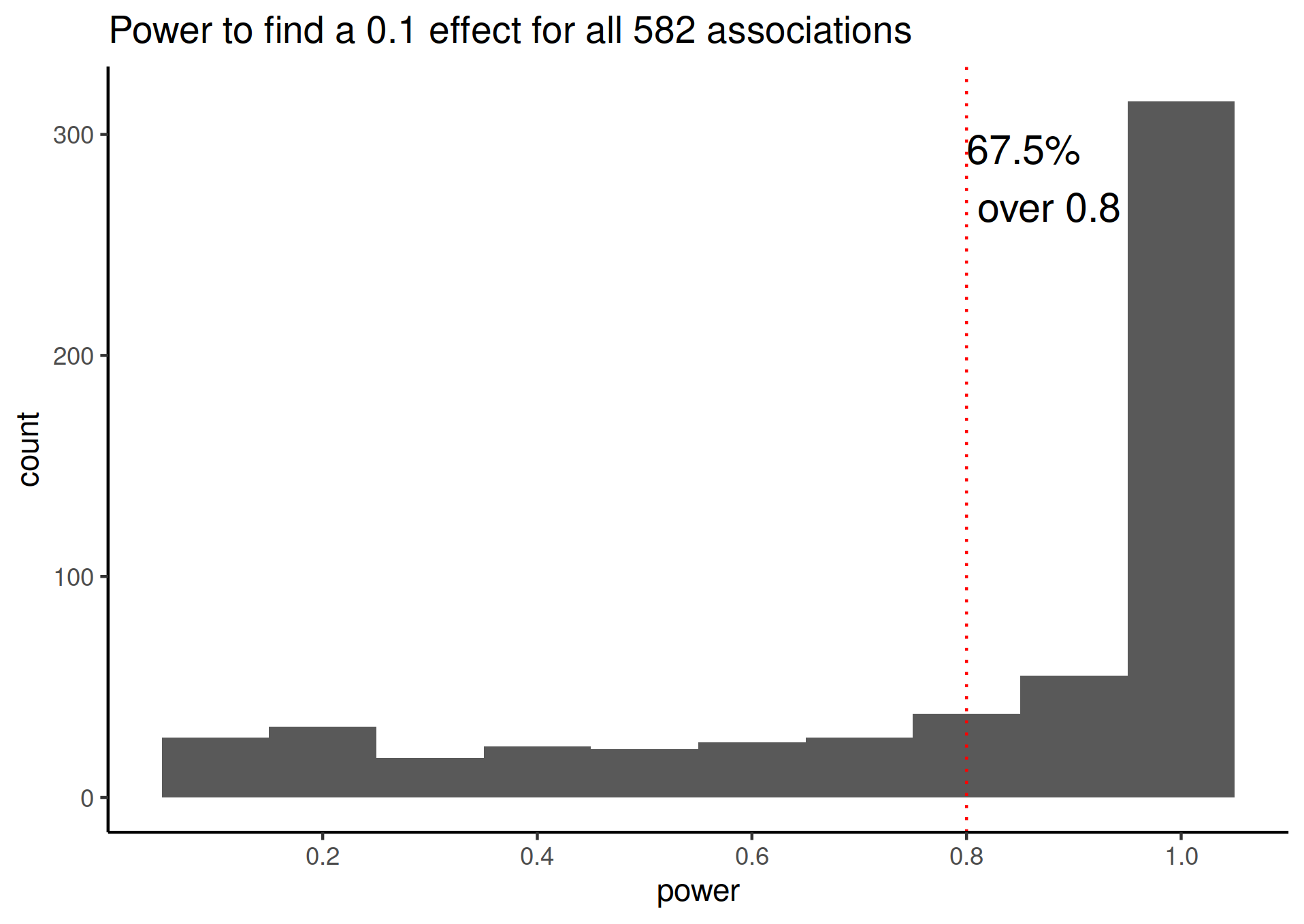

**Supplementary Figure 2. Histogramm of power for all evaluated associations. A 0.1 effect can be interpreted as a 1 SD change in exposure result in a ~10% change in the odds of contracting a disease.**

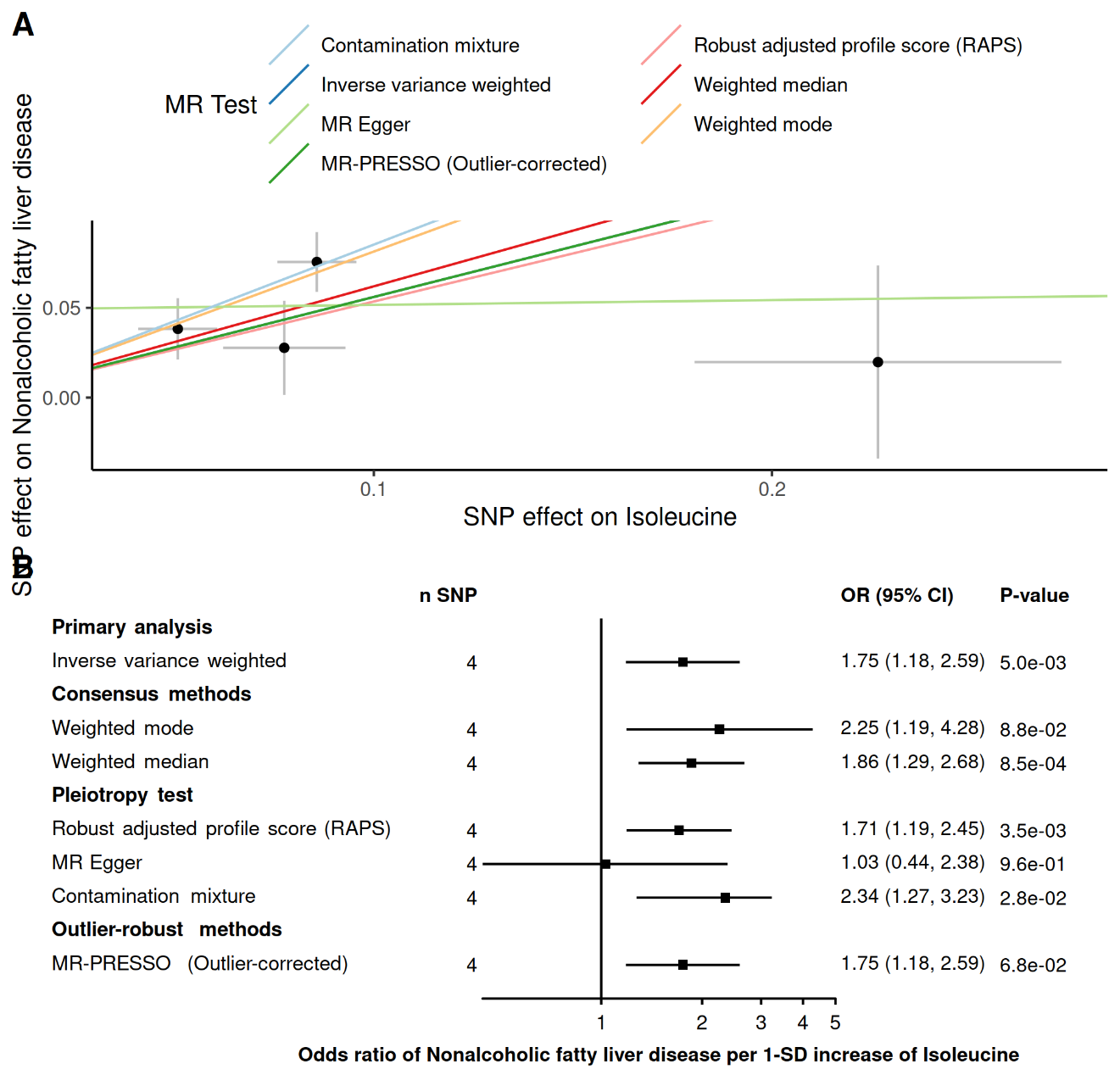

**Supplementary figure 3. Robust MR methods on the effect of Isoleucine on non-alcoholic fatty liver disease.** Panel A. Scatter plot of the associations where each dot represents one genetic instrument and each line represents the fitted estimates of different robust MR methods. Panel B. Forest plot of the association allowing to represent the uncertainty of the estimate.

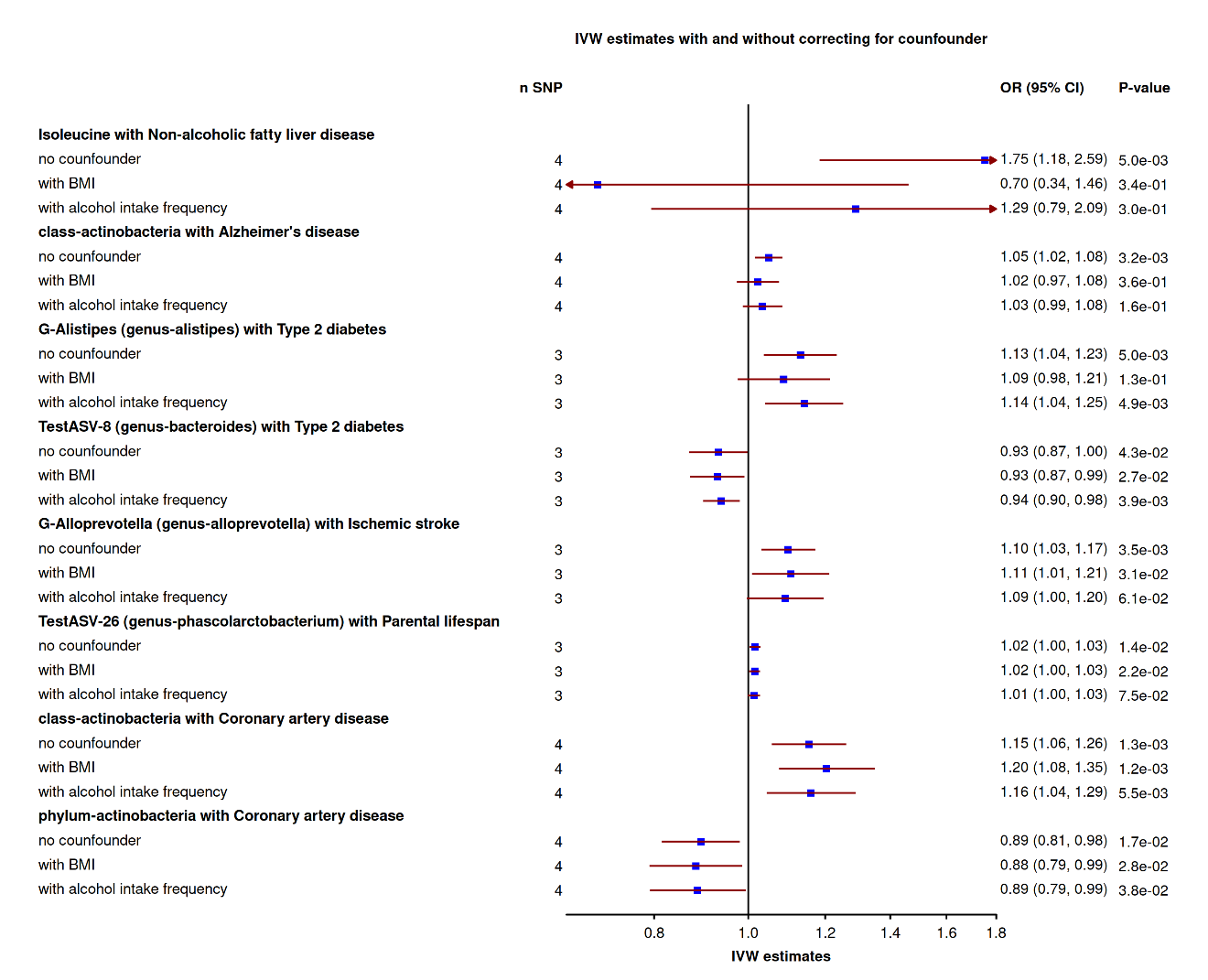

**Supplementary Figure 4. IVW-MR results before and after correcting for BMI and alcohol intake frequency using multivariable MR framework.**

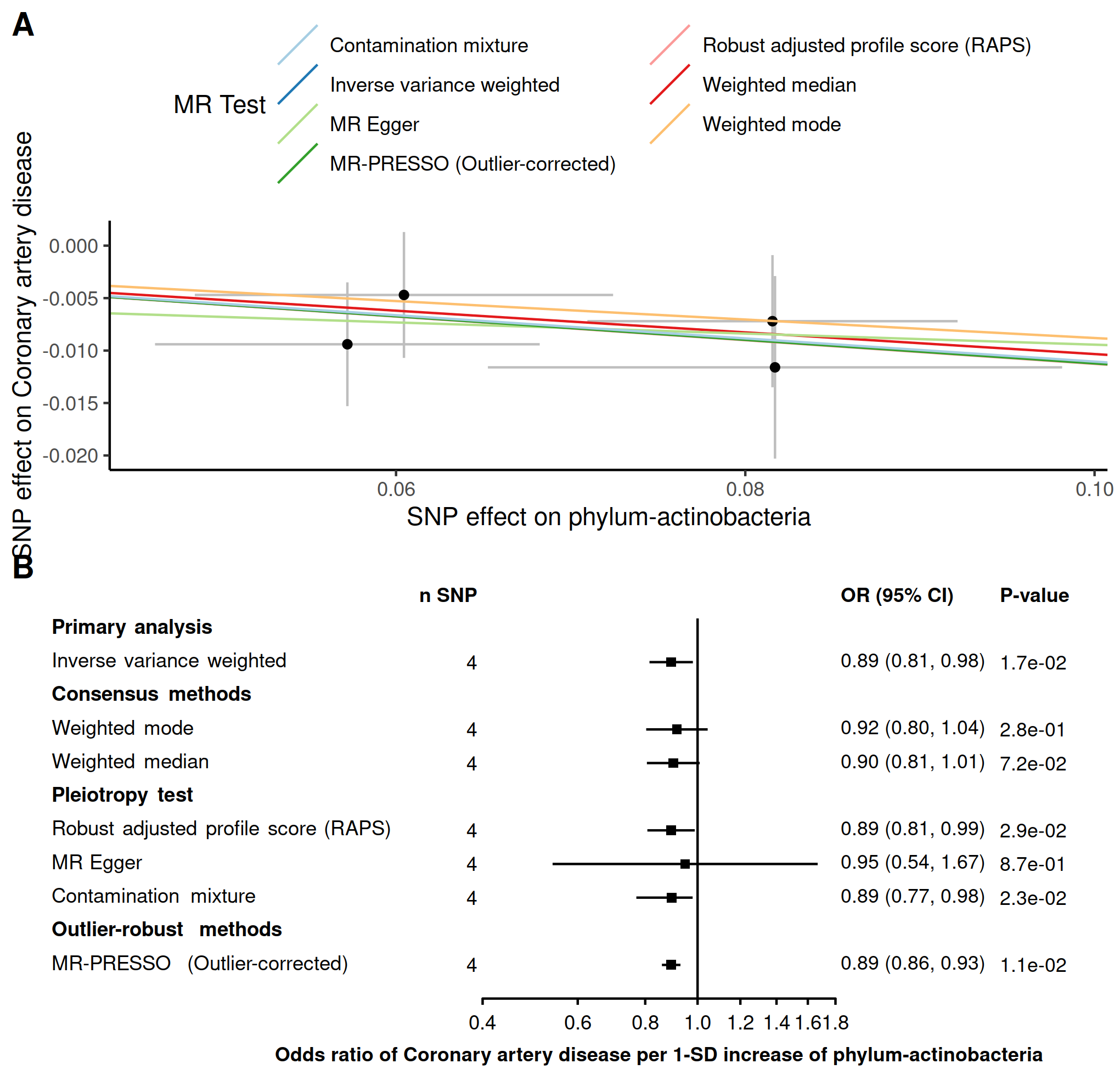

**Supplementary figure 5. Robust MR methods on the effect of Actinobacteria Phylum on coronary artery disease.** Panel A. Scatter plot of the associations where each dot represents one genetic instrument and each line represents the fitted estimates of different robust MR methods. Panel B. Forest plot of the association allowing to represent the uncertainty of the estimate.

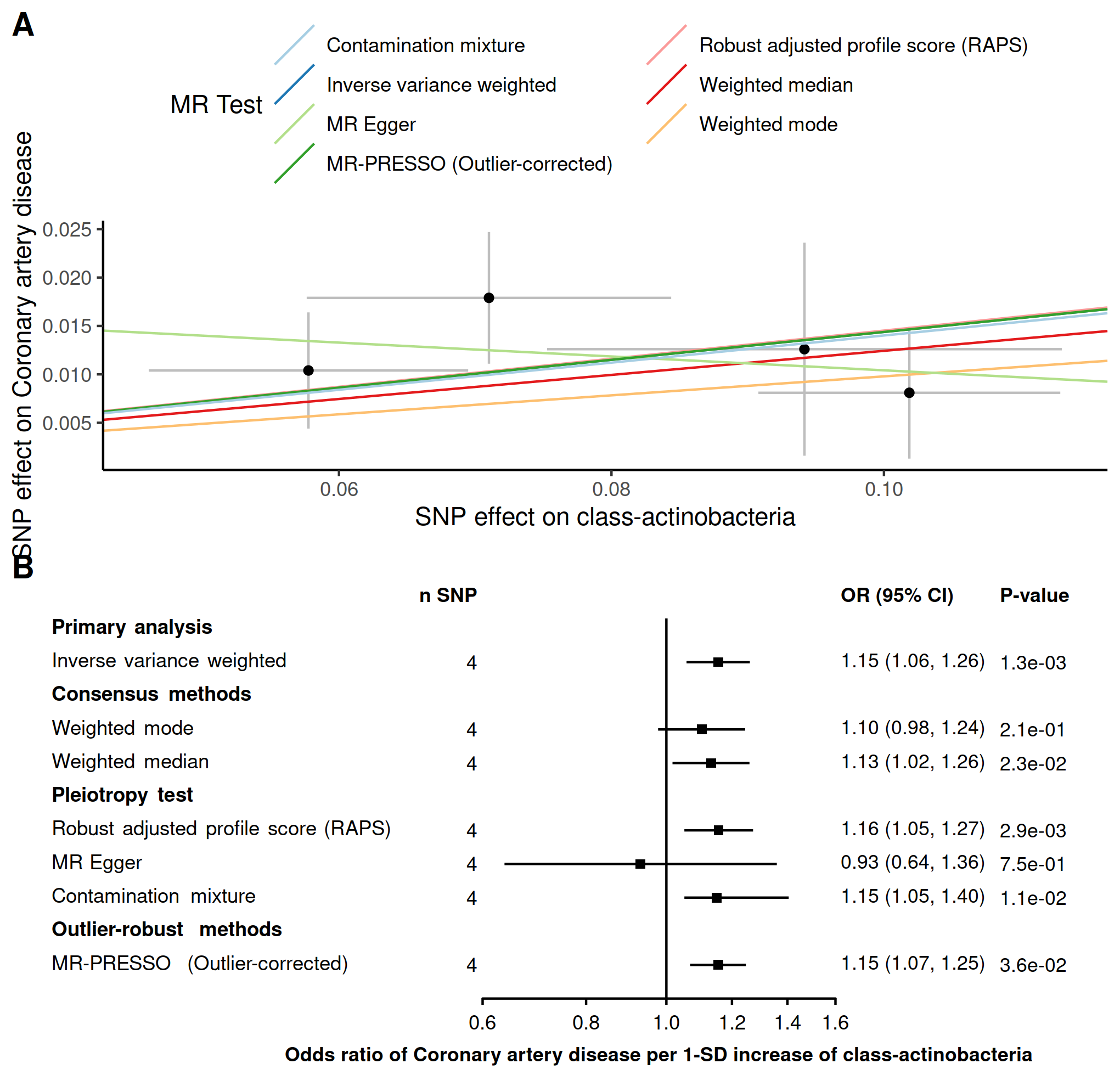

**Supplementary figure 6. Robust MR methods on the effect of *Actinobacteria* class on coronary artery disease.** Panel A. Scatter plot of the associations where each dot represents one genetic instrument and each line represents the fitted estimates of different robust MR methods. Panel B. Forest plot of the association allowing to represent the uncertainty of the estimate.

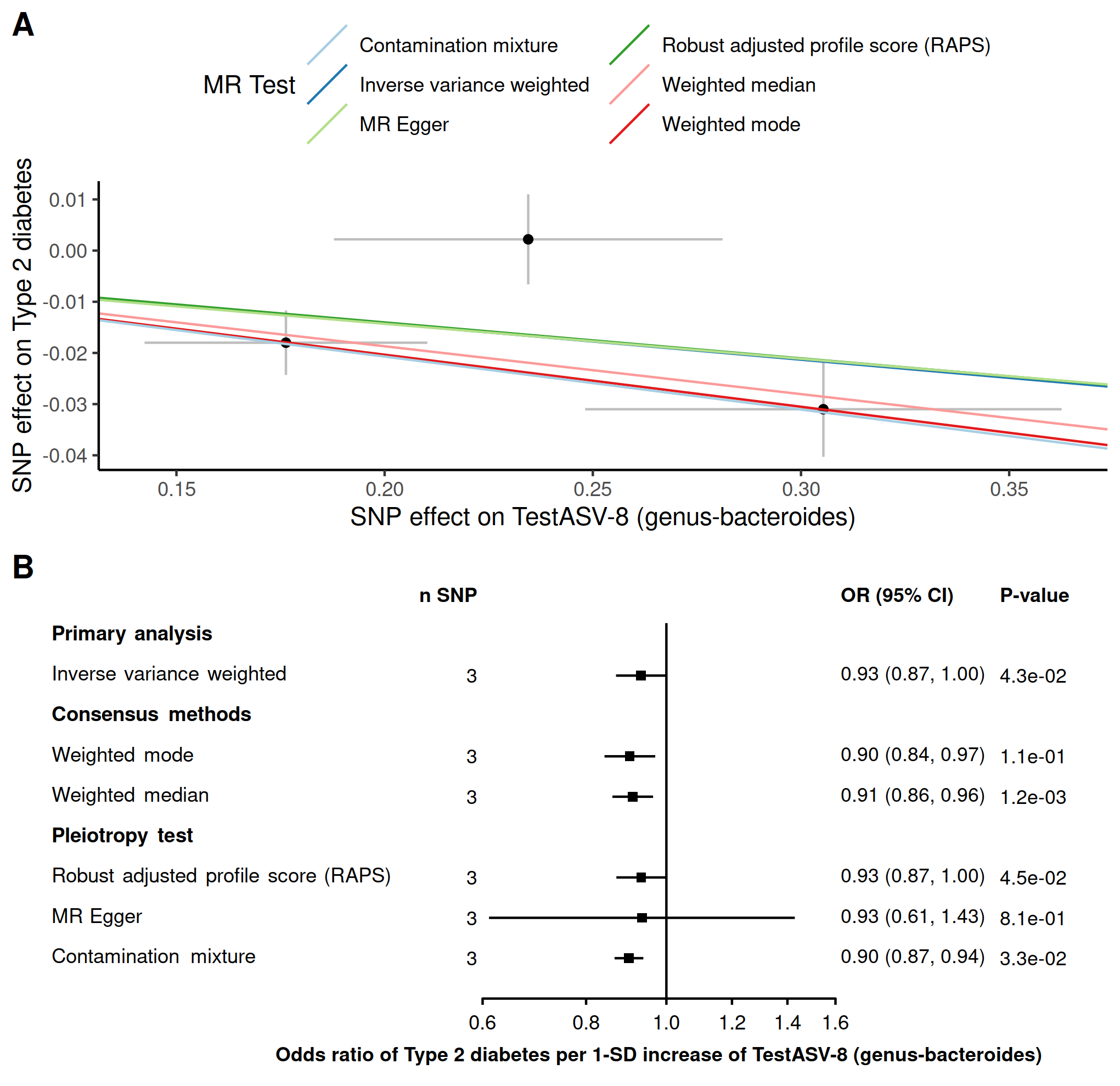

**Supplementary figure 7. Robust MR methods on the effect of *Bacteroides* genus on Type 2 diabetes.** Panel A. Scatter plot of the associations where each dot represents one genetic instrument and each line represents the fitted estimates of different robust MR methods. Panel B. Forest plot of the association allowing to represent the uncertainty of the estimate.

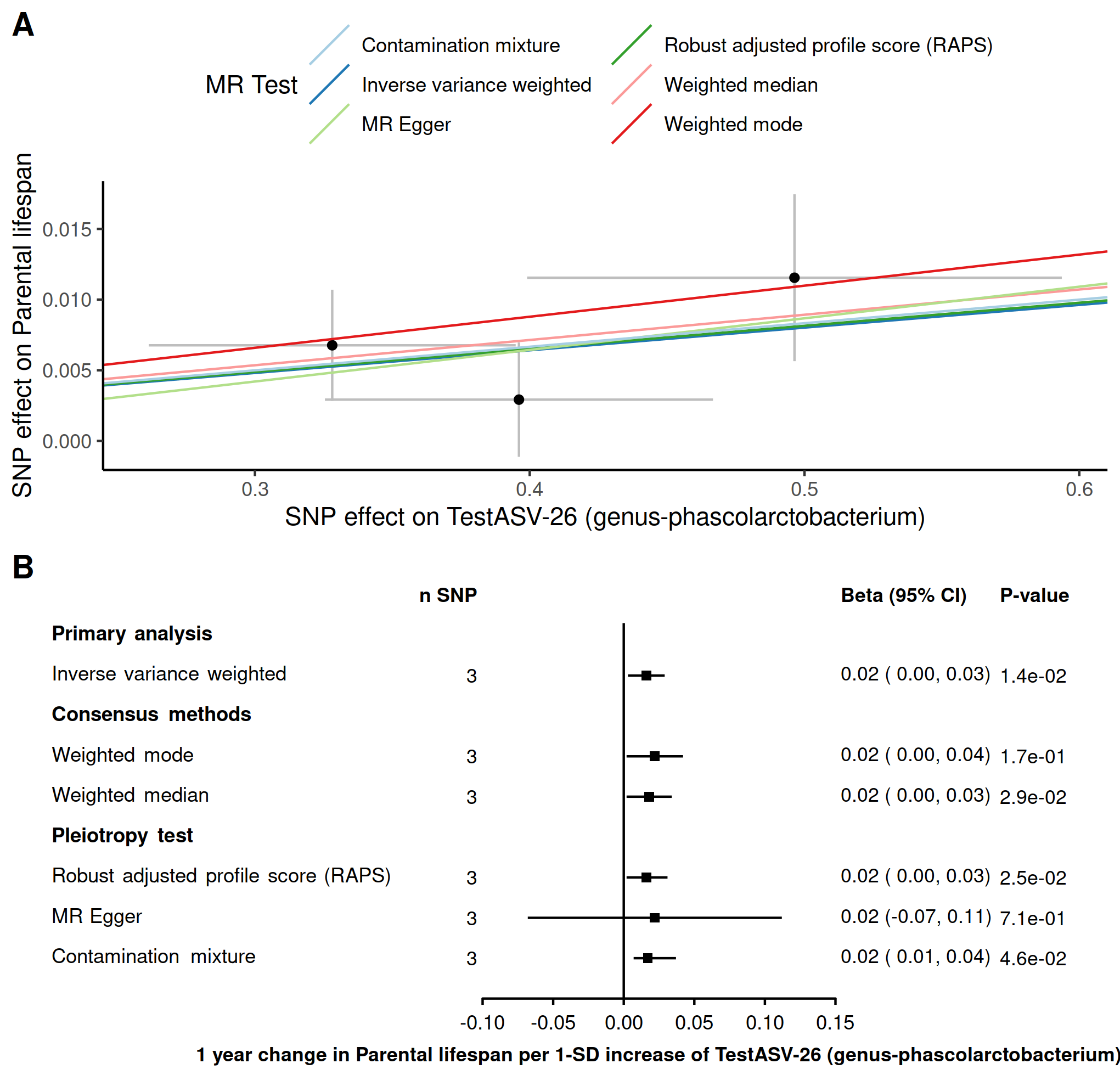

**Supplementary figure 8. Robust MR methods on the effect of *Phascolarctobacterium* genus on parental lifespan.** Panel A. Scatter plot of the associations where each dot represents one genetic instrument and each line represents the fitted estimates of different robust MR methods. Panel B. Forest plot of the association allowing to represent the uncertainty of the estimate.

**Supplementary Tables**

**Table 1. Robust MR methods for the association with inflammatory bowel disease (IBD) and all 10 health outomes.**

| **outcome** | **exposure** | **method** | **nsnp** | **b** | **se** | **pval** | **lci** | **uci** | **type_of_test** |
| --- | --- | --- | --- | --- | --- | --- | --- | --- | --- |
| Deelen_longevity | IBD | Inverse variance weighted | 106 | 0,003641 | 0,02046224 | 0,85875409 | -0,0364645 | 0,0437475 | Primary analysis |
| Deelen_longevity | IBD | Weighted mode | 106 | -0,006716 | 0,04410536 | 0,879255977 | -0,093163 | 0,07973 | Consensus methods |
| Deelen_longevity | IBD | Weighted median | 106 | -0,011062 | 0,03461518 | 0,749284107 | -0,0789082 | 0,0567833 | Consensus methods |
| Deelen_longevity | IBD | Robust adjusted profile score (RAPS) | 106 | -0,001818 | 0,02120221 | 0,931658774 | -0,0433746 | 0,0397381 | Pleiotropy test |
| Deelen_longevity | IBD | MR Egger | 106 | 0,056268 | 0,04877928 | 0,251344153 | -0,0393399 | 0,1518749 | Pleiotropy test |
| Deelen_longevity | IBD | MR-PRESSO (Outlier-corrected) | 106 | 0,003641 | 0,02046224 | 0,85909697 | -0,0364645 | 0,0437475 | Outlier-robust methods |
| Deelen_longevity | IBD | Contamination mixture | 106 | -0,023228 |  | 0,494876628 | -0,0732279 | 0,0567721 | Pleiotropy test |
| Howard_Depression | IBD | Inverse variance weighted | 106 | -0,000305 | 0,00584643 | 0,958443671 | -0,0117636 | 0,0111544 | Primary analysis |
| Howard_Depression | IBD | Weighted mode | 106 | -0,007165 | 0,01169076 | 0,541278505 | -0,030079 | 0,0157488 | Consensus methods |
| Howard_Depression | IBD | Weighted median | 106 | -0,005578 | 0,00743726 | 0,453274679 | -0,0201547 | 0,0089993 | Consensus methods |
| Howard_Depression | IBD | Robust adjusted profile score (RAPS) | 106 | 2,2E-05 | 0,00551073 | 0,996820674 | -0,0107791 | 0,010823 | Pleiotropy test |
| Howard_Depression | IBD | MR Egger | 106 | -0,01677 | 0,01411727 | 0,23757835 | -0,0444398 | 0,0108999 | Pleiotropy test |
| Howard_Depression | IBD | MR-PRESSO (Outlier-corrected) | 104 | 0,000326 | 0,00492175 | 0,94727894 | -0,0093204 | 0,0099729 | Outlier-robust methods |
| Howard_Depression | IBD | Contamination mixture | 106 | -0,00454 |  | 1 | -0,0145399 | 0,0154601 | Pleiotropy test |
| Jansen_Alzheimer | IBD | Inverse variance weighted | 106 | -0,00385 | 0,00285026 | 0,176760216 | -0,0094366 | 0,0017364 | Primary analysis |
| Jansen_Alzheimer | IBD | Weighted mode | 106 | -0,00726 | 0,00562871 | 0,199938016 | -0,0182925 | 0,0037721 | Consensus methods |
| Jansen_Alzheimer | IBD | Weighted median | 106 | -0,00507 | 0,00402428 | 0,207692408 | -0,0129579 | 0,0028173 | Consensus methods |
| Jansen_Alzheimer | IBD | Robust adjusted profile score (RAPS) | 106 | -0,003635 | 0,00289789 | 0,20967295 | -0,0093152 | 0,0020446 | Pleiotropy test |
| Jansen_Alzheimer | IBD | MR Egger | 106 | -0,004655 | 0,00697113 | 0,505796937 | -0,0183181 | 0,0090087 | Pleiotropy test |
| Jansen_Alzheimer | IBD | MR-PRESSO (Outlier-corrected) | 105 | -0,003234 | 0,00275484 | 0,243047735 | -0,0086339 | 0,0021651 | Outlier-robust methods |
| Jansen_Alzheimer | IBD | Contamination mixture | 106 | -0,007115 |  | 0,113236902 | -0,0071151 | -0,0071151 | Pleiotropy test |
| Mahajan_Type2diabetes | IBD | Inverse variance weighted | 106 | -0,00723 | 0,01483224 | 0,62592333 | -0,0363015 | 0,0218409 | Primary analysis |
| Mahajan_Type2diabetes | IBD | Weighted mode | 106 | -0,009227 | 0,01608152 | 0,567335917 | -0,0407472 | 0,0222923 | Consensus methods |
| Mahajan_Type2diabetes | IBD | Weighted median | 106 | -0,001982 | 0,01228976 | 0,871852694 | -0,0260703 | 0,0221055 | Consensus methods |
| Mahajan_Type2diabetes | IBD | Robust adjusted profile score (RAPS) | 106 | 0,0042 | 0,01263125 | 0,739494568 | -0,0205571 | 0,0289574 | Pleiotropy test |
| Mahajan_Type2diabetes | IBD | MR Egger | 106 | -0,035039 | 0,03621196 | 0,335485351 | -0,1060144 | 0,0359365 | Pleiotropy test |
| Mahajan_Type2diabetes | IBD | MR-PRESSO (Outlier-corrected) | 97 | 0,007944 | 0,00986209 | 0,422499036 | -0,0113854 | 0,027274 | Outlier-robust methods |
| Mahajan_Type2diabetes | IBD | Contamination mixture | 106 | 0,000892 |  | 1 | -0,0191078 | 0,0208922 | Pleiotropy test |
| Malik_Stroke | IBD | Inverse variance weighted | 105 | -0,000266 | 0,01271229 | 0,983301061 | -0,0251822 | 0,02465 | Primary analysis |
| Malik_Stroke | IBD | Weighted mode | 105 | -0,015778 | 0,02481476 | 0,526293549 | -0,0644145 | 0,0328593 | Consensus methods |
| Malik_Stroke | IBD | Weighted median | 105 | -0,006893 | 0,01863445 | 0,711467858 | -0,0434161 | 0,0296309 | Consensus methods |
| Malik_Stroke | IBD | Robust adjusted profile score (RAPS) | 105 | -0,004541 | 0,01375722 | 0,741363992 | -0,0315047 | 0,0224236 | Pleiotropy test |
| Malik_Stroke | IBD | MR Egger | 105 | -0,063892 | 0,03013486 | 0,036391269 | -0,1229562 | -0,0048275 | Pleiotropy test |
| Malik_Stroke | IBD | Contamination mixture | 105 | -0,037149 |  | 0,091862543 | -0,0571493 | 0,0028507 | Pleiotropy test |
| NAFLD | IBD | Inverse variance weighted | 105 | 0,039661 | 0,01757846 | 0,024056144 | 0,00520729 | 0,0741148 | Primary analysis |
| NAFLD | IBD | Weighted mode | 105 | 0,019721 | 0,03864586 | 0,610916229 | -0,0560245 | 0,0954672 | Consensus methods |
| NAFLD | IBD | Weighted median | 105 | 0,037696 | 0,02815506 | 0,180609328 | -0,0174876 | 0,0928802 | Consensus methods |
| NAFLD | IBD | Robust adjusted profile score (RAPS) | 105 | 0,035203 | 0,01766296 | 0,046258509 | 0,00058335 | 0,0698222 | Pleiotropy test |
| NAFLD | IBD | MR Egger | 105 | 0,003657 | 0,04320456 | 0,932709438 | -0,081024 | 0,0883379 | Pleiotropy test |
| NAFLD | IBD | MR-PRESSO (Outlier-corrected) | 105 | 0,039661 | 0,01757846 | 0,026148469 | 0,00520729 | 0,0741148 | Outlier-robust methods |
| NAFLD | IBD | Contamination mixture | 105 | 0,048141 |  | 0,2871298 | -0,0318595 | 0,0881405 | Pleiotropy test |
| Timmers_parental_lifespan | IBD | Inverse variance weighted | 105 | -0,011097 | 0,00583417 | 0,057155189 | -0,0225323 | 0,0003377 | Primary analysis |
| Timmers_parental_lifespan | IBD | Weighted mode | 105 | 0,004666 | 0,01030951 | 0,651768399 | -0,0155404 | 0,0248729 | Consensus methods |
| Timmers_parental_lifespan | IBD | Weighted median | 105 | -0,002377 | 0,00716252 | 0,739966134 | -0,0164158 | 0,0116613 | Consensus methods |
| Timmers_parental_lifespan | IBD | Robust adjusted profile score (RAPS) | 105 | -0,009462 | 0,00529723 | 0,074069015 | -0,0198444 | 0,0009207 | Pleiotropy test |
| Timmers_parental_lifespan | IBD | MR Egger | 105 | 0,010889 | 0,01401043 | 0,438805036 | -0,0165711 | 0,0383498 | Pleiotropy test |
| Timmers_parental_lifespan | IBD | MR-PRESSO (Outlier-corrected) | 102 | -0,010117 | 0,00485283 | 0,039606153 | -0,0196289 | -0,0006058 | Outlier-robust methods |
| Timmers_parental_lifespan | IBD | Contamination mixture | 105 | 9,99E-05 |  | 0,889404133 | -0,0099001 | 0,0100999 | Pleiotropy test |
| ukb-b-12141_osteoporosis | IBD | Inverse variance weighted | 104 | 0,000555 | 0,00032743 | 0,089825391 | -8,634E-05 | 0,0011972 | Primary analysis |
| ukb-b-12141_osteoporosis | IBD | Weighted mode | 104 | -0,000576 | 0,00059201 | 0,33268586 | -0,0017365 | 0,0005841 | Consensus methods |
| ukb-b-12141_osteoporosis | IBD | Weighted median | 104 | -0,000299 | 0,00044513 | 0,501305983 | -0,0011718 | 0,0005731 | Consensus methods |
| ukb-b-12141_osteoporosis | IBD | Robust adjusted profile score (RAPS) | 104 | 0,000383 | 0,00033623 | 0,254611813 | -0,000276 | 0,0010421 | Pleiotropy test |
| ukb-b-12141_osteoporosis | IBD | MR Egger | 104 | -0,000704 | 0,00078793 | 0,373964753 | -0,0022479 | 0,0008407 | Pleiotropy test |
| ukb-b-12141_osteoporosis | IBD | MR-PRESSO (Outlier-corrected) | 103 | 0,00049 | 0,00031312 | 0,12068399 | -0,0001237 | 0,0011037 | Outlier-robust methods |
| ukb-b-12141_osteoporosis | IBD | Contamination mixture | 104 | -0,003043 |  | 1 | -0,0030428 | -0,0030428 | Pleiotropy test |
| van_der_Harst_CAD | IBD | Inverse variance weighted | 106 | -0,003883 | 0,01149277 | 0,735435659 | -0,0264093 | 0,0186424 | Primary analysis |
| van_der_Harst_CAD | IBD | Weighted mode | 106 | -0,009808 | 0,01345536 | 0,467653609 | -0,0361808 | 0,0165642 | Consensus methods |
| van_der_Harst_CAD | IBD | Weighted median | 106 | -0,004037 | 0,00963891 | 0,675360811 | -0,0229291 | 0,0148555 | Consensus methods |
| van_der_Harst_CAD | IBD | Robust adjusted profile score (RAPS) | 106 | -0,000342 | 0,00931091 | 0,970667268 | -0,0185918 | 0,017907 | Pleiotropy test |
| van_der_Harst_CAD | IBD | MR Egger | 106 | -0,051225 | 0,02786987 | 0,068916485 | -0,1058498 | 0,0034001 | Pleiotropy test |
| van_der_Harst_CAD | IBD | MR-PRESSO (Outlier-corrected) | 99 | 0,000492 | 0,00752468 | 0,948046846 | -0,0142568 | 0,0152399 | Outlier-robust methods |
| van_der_Harst_CAD | IBD | Contamination mixture | 106 | 0,010575 |  | 0,735525598 | -0,0094248 | 0,0205752 | Pleiotropy test |
| Wuttke_Chronic_kidney | IBD | Inverse variance weighted | 106 | 0,017702 | 0,01409906 | 0,209293712 | -0,0099326 | 0,0453357 | Primary analysis |
| Wuttke_Chronic_kidney | IBD | Weighted mode | 106 | -0,017729 | 0,02425492 | 0,466429351 | -0,0652691 | 0,0298102 | Consensus methods |
| Wuttke_Chronic_kidney | IBD | Weighted median | 106 | 0,000505 | 0,01633164 | 0,975348902 | -0,0315054 | 0,0325147 | Consensus methods |
| Wuttke_Chronic_kidney | IBD | Robust adjusted profile score (RAPS) | 106 | 0,016517 | 0,01405239 | 0,239853101 | -0,0110262 | 0,0440592 | Pleiotropy test |
| Wuttke_Chronic_kidney | IBD | MR Egger | 106 | -0,021135 | 0,03425141 | 0,538555966 | -0,0882673 | 0,0459982 | Pleiotropy test |
| Wuttke_Chronic_kidney | IBD | MR-PRESSO (Outlier-corrected) | 103 | 0,0142 | 0,01187702 | 0,234640225 | -0,0090793 | 0,0374787 | Outlier-robust methods |
| Wuttke_Chronic_kidney | IBD | Contamination mixture | 106 | 0,009855 |  | 0,510854467 | -0,0301447 | 0,0498553 | Pleiotropy test |

**Supplementary Table 2. Information on exposures GWAS data.**

| **exposures raw data** | **p** | **r^2** | **window** | **link to find the data** | **other information to find the data** |
| --- | --- | --- | --- | --- | --- |
| Liu-IBD | 5x10^-8 of a related diseases | did not clump | did not clump | <https://www.nature.com/articles/ng.3359#Sec17> | Supplementary Table 1 sheet 3. |
| Sanna | P < 1 × 10−5 | 0.001 | 10000 | <https://static-content.springer.com/esm/art%3A10.1038%2Fs41588-019-0350-x/MediaObjects/41588_2019_350_MOESM1_ESM.pdf> | for propionate Supplementary Table 8. for PWY-5022 supplementary table 4 |
| framingham | P < 5 × 10−5 | did not clump | did not clump | <https://www.cell.com/action/showFullTableHTML?isHtml=true&tableId=tbl2&pii=S1550-4131%2813%2900257-X> | Table 2 Genome-wide Significant Loci Associated with Metabolites |
| kettunen | all | all | all | <http://www.computationalmedicine.fi/data#NMR_GWAS> | The four datasets downloaded had suffix c("Ile", "Leu", "Val", "Ace") for isoleucine, leucine, valine and acetate respectively |
| Qin | 5*10^-8 | independent hits |  | <https://www.medrxiv.org/content/10.1101/2020.09.12.20193045v1> | Table S1: 583 genome-wide significant associations between bacterial taxa abundances and genetic variants. |
| Lopera-Maya | 5*10^-8 | 0.5 | 10000 | <https://www.biorxiv.org/content/10.1101/2020.12.09.417642v1.supplementary-material> | Supplementary Table 1. Genome-wide significant results |
| ruhlemann | max pvalue = 7.91e-06 | 10000 most significant hit. | did not clump | <https://www.nature.com/articles/s41588-020-00747-1> | Supplementary_Table_S3_Abundance |
| kurilshikov | 1*10^-6 | did not clump | did not clump | <https://www.nature.com/articles/s41588-020-00763-1#Sec26> | suppelmentary table 7 |
| IBD for the LD score by Liu | filteredI NFO>0.3 and with more than 2 studies/datasets | did not clump | did not clump | <https://www.ibdgenetics.org/downloads.html> | [Latest combined GWAS and Immunochip trans-ancestry summary statistics](ftp://ftp.sanger.ac.uk/pub/consortia/ibdgenetics/iibdgc-trans-ancestry-filtered-summary-stats.tgz) |

**Supplementary Table 3. Information on outcomes GWAS data.**

| **outcomes** | **p** | **r^2** | **info** | **link to find the data** | **other information to find the data** |
| --- | --- | --- | --- | --- | --- |
| Coronary heart disease | all | all | European | <https://data.mendeley.com/datasets/gbbsrpx6bs/1> | <https://www.cardiomics.net/download-data> |
| Ischemic stroke | all | all | European | <https://www.megastroke.org/download.html> |  |
| Alzheimer disease | all | all | European | <https://ctg.cncr.nl/software/summary_statistics> | [AD_sumstats_Jansenetal_2019sept.txt.gz](https://ctg.cncr.nl/documents/p1651/AD_sumstats_Jansenetal_2019sept.txt.gz) |
| Depression | all | all | contains UKB but not 23 and me | <https://datashare.is.ed.ac.uk/handle/10283/3203> | [Genome-wide summary statistics from a meta-analysis of PGC and UK Biobank](https://datashare.is.ed.ac.uk/bitstream/handle/10283/3203/PGC_UKB_depression_genome-wide.txt?sequence=3&isAllowed=y) |
| Osteoporosis | all | all | UKB accesbile via ieugwasr package | We obtained the summary statistics via the ieugwasr package (Hemani et al., 2018). | id = ukb-b-12141 |
| Non-alcoholic fatty liver disease (NAFLD) |  |  | GWAS by our group | not yet publically accessible |  |
| Type-2 diabetes | all | all | only European | <http://diagram-consortium.org/downloads.html> | T2D GWAS meta-analysis - Unadjusted for BMI Published in in Mahajan et al (2018b) |
| Chronic Kidney disease | all | all | only European, CKD =Chronik Kidney Disease | http://ckdgen.imbi.uni-freiburg.de/ | [CKD overall European ancestry](http://ckdgen.imbi.uni-freiburg.de/files/Wuttke2019/CKD_overall_EA_JW_20180223_nstud23.dbgap.txt.gz) |
| Parental Lifespan | all | all | only european ancestry Continuous measure in years. | <https://datashare.is.ed.ac.uk/handle/10283/3209> | [lifegen_phase2_bothpl_alldr_2017_09_18.tsv.gz (461.0Mb)](https://datashare.is.ed.ac.uk/bitstream/handle/10283/3209/lifegen_phase2_bothpl_alldr_2017_09_18.tsv.gz?sequence=1&isAllowed=y) |
| Longevity | all | all | only european | <https://www.longevitygenomics.org/downloads> | [90th percentile cases vs all controls](https://www.ebi.ac.uk/gwas/studies/GCST008598) |
| osteoporosis | all | all | only european. | http://www.gefos.org/?q=content/data-release-2018 | [Morrisetal2018.NatGen.SumStats.tar.gz](http://www.gefos.org/sites/default/files/Morrisetal2018.NatGen.SumStats.tar_0.gz) |
